# Transferability and accuracy of electronic health record-based predictors compared to polygenic scores

**DOI:** 10.1101/2024.10.08.24315073

**Authors:** Kira E. Detrois, Tuomo Hartonen, Maris Teder-Laving, Bradley Jermy, Kristi Läll, Zhiyu Yang, Estonian Biobank research team, FinnGen, Reedik Mägi, Samuli Ripatti, Andrea Ganna

**Affiliations:** Institute for Molecular Medicine Finland, FIMM, HiLIFE, University of Helsinki, Helsinki, Finland; Estonian Genome Centre, Institute of Genomics, University of Tartu, Tartu, Estonia; Broad Institute of MIT and Harvard, Cambridge, MA, USA; Analytic and Translational Genetics Unit, Massachusetts General Hospital, Boston, MA, USA; Department of Public Health, University of Helsinki, Helsinki, Finland

## Abstract

Electronic health record (EHR)-based phenotype risk scores (PheRS) leverage individuals’ health trajectories to infer disease risk. Similarly, polygenic scores (PGS) use genetic information to estimate disease risk. While PGS generalizability has been previously studied, less is known about PheRS transferability across healthcare systems and whether PheRS provide complementary risk information to PGS.

We trained PheRS to predict the onset of 13 common diseases with high health burden in a total of 845,929 individuals (age 32-70) from 3 biobank-based studies from Finland (FinnGen), the UK (UKB) and Estonia (EstB). The PheRS were based on elastic-net models, incorporating up to 242 diagnoses captured in the EHR up to 10 years before baseline. Individuals were followed up for a maximum of 8 years, during which disease incidence was observed. PGS were calculated for each disease using recent publicly available results from genome-wide association studies.

All 13 PheRS were significantly associated with the diseases of interest. The PheRS trained in different biobanks utilized partially distinct diagnoses, reflecting differences in medical code usage across the countries. Even with the large variability in the prevalence of various diagnoses, most PheRS trained in the UKB or EstB transferred well to FinnGen without re-training. PheRS and PGS were only moderately correlated (Pearson’s *r* ranging from 0.00 to 0.08), and models including both PheRS and PGS improved onset prediction compared to PGS alone for 8/13 diseases. PheRS was able to identify a subset of individuals at high-risk better than PGS for 8/13 disease.

Our results indicate that EHR-based risk scores and PGS capture largely independent information and provide additive benefits for disease risk prediction. Furthermore, for many diseases the PheRS models transfer well between different EHRs. Given the large availability of EHR, PheRS can provide a complementary tool to PGS for risk stratification.

## Introduction

With the advent of large-scale genetic studies and the widespread availability of electronic health record (EHR) data, it is possible to combine these resources to more efficiently predict the risk of a wide range of diseases^1–3^. Disease risk estimation can guide the efficient allocation of screening, preventative interventions, and treatments in the early stages of diseases. Two lines of research have emerged in the past years. Some researchers have focused on machine learning approaches for EHR data ^2,4^ and showed some promising results in deriving EHR-based predictors for pancreatic cancer^5^ and cardiovascular disease^6–8^, among others. Many studies have focused on genetic data. Polygenic scores (PGSs) use combined information from a person’s genome to estimate their genetic risk of developing a specific disease or trait. Numerous studies have examined the predictive ability of PGS across multiple diseases, and there is an extensive discussion about their clinical and public health value^9–24^.

EHR and PGS-based prediction models have different strengths and limitations. EHRs allow access to a vast variety of data, including but not limited to disease diagnosis history, laboratory measurements, free text reports, and various socio-economic information^25^. However, EHRs are also known to be noisy^1,2^, and the models are expected to suffer from poor generalizability because of differences in data availability, as well as in clinical and recording practices across healthcare systems^2,3,5,25–28^. So far, most research has been conducted on a single EHR with limited work on validating the models in different EHR systems and countries^2,27,29,30^. Recent studies, however, show promising results when validating EHR-based predictors in a different healthcare system in the US and UK. For example, an EHR-based prediction model trained in a US study (*BioMe*) outperformed conventional clinical guidelines in predicting coronary artery disease (CAD) susceptibility, and the results could be externally replicated in the UK Biobank (UKB)^7,8^. A similar recent study successfully transferred an EHR-based model trained in the *BioMe* study for the prediction of autoimmune diseases to All of US, another US-based study. Another systematic effort to train deep learning-based prediction models on the UKB EHR data for 1.568 diseases showed that when transferring these models to the All of Us study, 1.347 (85.9%) of the models improved disease onset prediction over a baseline model with age and sex^31^.

PGS are less likely to suffer from measurement errors compared to EHR-based models, however, they are known to be poorly transferable across ancestries, thus risking increasing health disparities^14,32^. PGS are also not routinely measured in the healthcare setting, although some healthcare systems have piloted programs to return PGS to individuals^33,34^. Further, as PGS keep improving through larger and more representative genome-wide association studies, there is a growing interest in the integration of other predictors and risk factors to better capture the disease risk of individuals. Some studies have been recently published integrating, for example, proteomics^35,36^ or metabolomics-based risk scores^37,38^ with PGS. Compared to omics, EHR data has the advantage that it is already routinely electronically collected in many countries and does not require invasive and often relatively expensive additional measurements^3^. Importantly, there is a gap in our understanding of how PGS complement both established clinical risk factors and EHR-based risk scores. Numerous studies have investigated the additive value of PGS with clinical risk factors for a subset of diseases, including type 2 diabetes (T2D) and CAD^9,16,39^. For EHR-based risk scores and many other diseases the added benefit of PGS for disease onset prediction and risk stratification remains understudied.

In this study, we aimed to directly compare, within and across studies, the predictive performance and transferability of EHR-based scores *vs.* PGS using a longitudinal prospective design. We conducted this comparison across 13 common diseases and 3 large biobank-based studies with high-quality EHR: UK Biobank^40^ (UKB, United Kingdom), FinnGen^41^ (Finland) and Estonian Biobank (EstB, Estonia)^34^. We created the EHR-based scores using the PheRS (Phenotype Risk Score) framework^42,43^ with PheRS derived from longitudinal diagnostic codes translated into consistent disease diagnoses using phecodes^44^.

## Results

### Study overview

We included 845,929 individuals (Supplement Table 1) aged 32 to 70 on 01/01/2011 (**Figure 1A**). These individuals belong to 3 biobank-based studies (FinnGen, UKB, EstB) linked with national registers or EHRs. The individuals gathered a total of 293,019 new diagnoses during an 8-year prediction period (01/01/2011 – 31/12/2018) across 13 common and high-burden diseases: prostate cancer, breast cancer, colorectal cancer, lung cancer, type 2 diabetes (T2D), atrial fibrillation (AF), major depression (MDD), coronary heart disease (CHD), hip osteoarthritis (hip OA), knee osteoarthritis (knee OA), asthma, gout, and epilepsy. We observed the highest number of events for knee OA (N=43,767) and the lowest for lung cancer (N=4,796, **Figure 1C**, Supplement Table 2).

**Figure 1:**
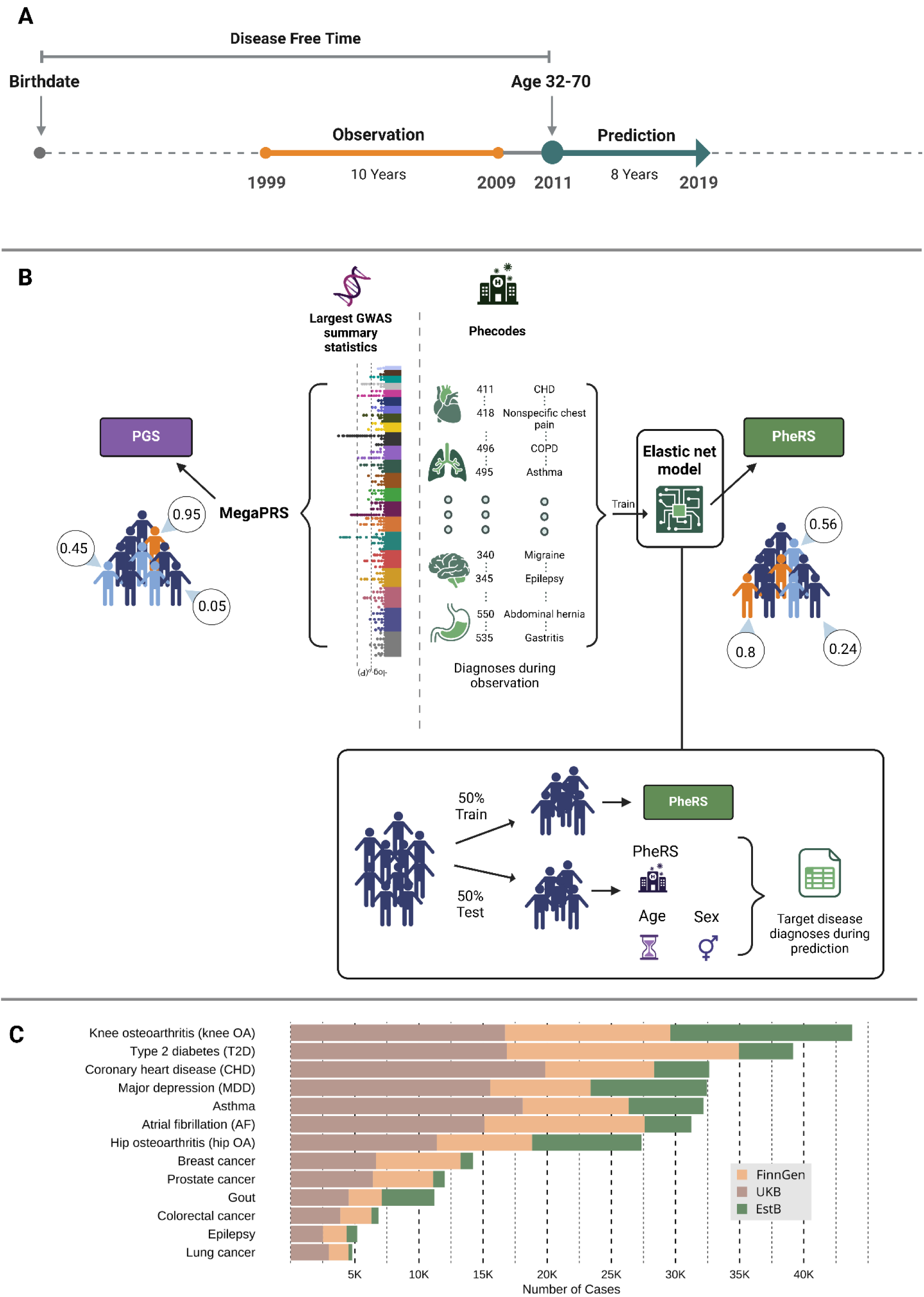
Study overview. **Panel A** Outline of the study design. A separate study is conducted for each of the 13 diseases in the three biobank-based studies. Each study consists of an observation and a prediction period, separated by a 2-year washout period. Each disease’s case and control definitions were based on diagnoses acquired in the prediction period (1/1/2011 – 31/12/2018). We removed all individuals diagnosed before our baseline (1/1/2011) and only considered adults aged 32-70 in 2011 (see Methods for more details). **Panel B:** We compared the PGSs with PheRSs – trained on phecodes recorded during the observation period (1/1/1999 – 31/12/2008). The PGS were based on recent publicly available GWAS summary statistics using MegaPRS. Ultimately, each individual was assigned 13 different PGS and PheRS scores describing their risk of getting a disease diagnosis during the prediction period. We trained the PheRS on 50% of individuals separately in the three studies (FinnGen, UKB, EstB). In each study, we then used the other half of the population as a test set where we used the scores as predictors in Cox-proportional hazards models^45^ (Cox-PH). **Panel C**: Number of new diagnoses for each disease during the prediction period (1/1/2011 – 31/12/2018) for each of the 13 diseases in the three cohorts (green: EstB, yellow: FinnGen, brown: UKB). This figure was created with the help of BioRender.com.

### Construction of PGS and PheRS

We constructed the PGS and PheRS separately for each disease (**Figure 1B**). PGS were previously derived by the INTERVENE consortium^46^. PheRS were based on phecodes^47^ recorded during a 10-year observation period (01/01/1999 – 31/12/2009; **Figure 1A**), separated from the prediction period (01/01/2011 – 31/12/2018) by a 2-year washout period. In total, we considered 242 phecodes with a prevalence of at least 1% in any study. Each PheRS model was trained separately to predict disease occurrence in the prediction period using 50% of the individuals in each study. We used an elastic net model, a type of regularized regression method that combines the properties of both Ridge (L2) and Lasso (L1) regression^43^. The effect of age, sex and the ten first genetic principal components (PCs) were regressed out from both PheRS and PGS to make the scores comparable. A more detailed description of the PheRS construction can be found in the Methods. Disease prevalence during the prediction period (01/01/2011 – 31/12/2018, **Figure 1C**) varied substantially across the 3 studies. For example, we found a higher prevalence of knee OA in the EstB (12.3%, N=14,180) compared to FinnGen (4.8%, N=12,874) and the UKB (3.6%, N=16,713), while T2D diagnoses show a lower prevalence both in the EstB (3.6%, N=104,161) and UKB (3.6%, N=16,850) compared to FinnGen (6.8%, N=18,099; Supplement Table 2).

### PheRS were significantly associated with all 13 diseases

We evaluated the association between PheRS and 13 diseases independently from age and sex using Cox proportional hazard models (Cox-PH) on a test set in each study. All PheRS were significantly associated (p<0.05) with higher disease risk (**Figure 2A**, Supplement Table 3) with the largest association for gout (meta-analyzed hazard ratio (HR) per 1 standard deviation (SD) of PheRS=1.55; 95% confidence interval (CI): 1.43-1.67), T2D (HR=1.47; 95% CI: 1.36-1.59), and lung cancer (HR=1.47; 95% CI: 1.39-1.55). Further, adding the PheRS to a baseline model with age and sex significantly (p<0.05; two-tailed p-values based on the z-scores of the c-index differences) improved the predictive accuracy (c-index) in all three studies for 7/13 diseases: asthma, MDD, T2D, knee OA, hip OA, gout, and AF (**Figure 2B**, Supplement Figure 1A-1, Supplement Table 4). The improvement persisted for 4 of these diseases (asthma, MDD, T2D, knee OA) in all of the three studies when compared to a baseline including additionally highest achieved education level and the Charlson comorbidity index^48,49^ (CCI; Supplement Figure 1A-2). Overall, integrating education and CCI only led to minor improvements in the model’s discriminative ability (Supplement Figures 2&3).

**Figure 2:**
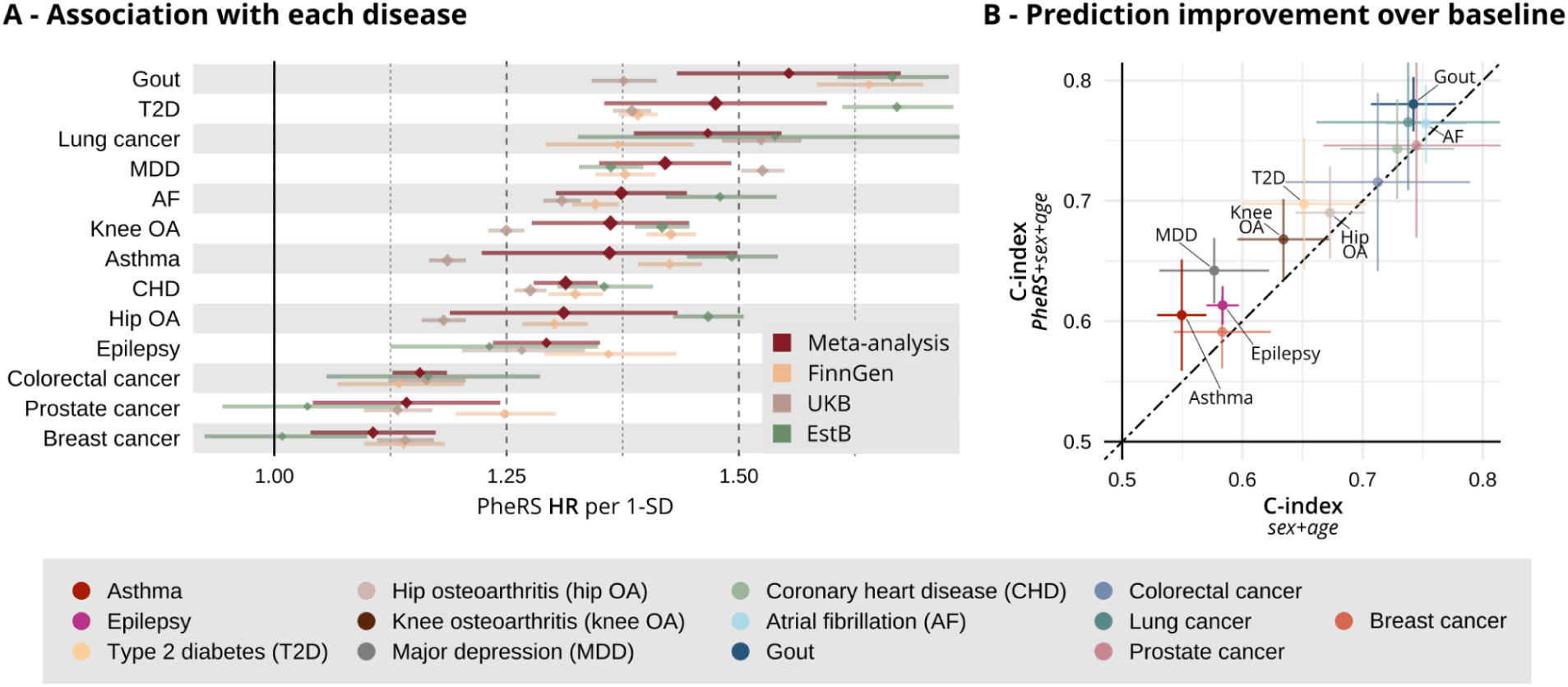
PheRS performance across studies. **Panel A**: Association between PheRS and disease onset during the prediction period independent of age and sex. The HRs and 95% CIs in each study – FinnGen: yellow, UKB: brown, EstB: green – and meta-analyzed results (red). The HRs are shown for an increase of the PheRS by 1 standard deviation (SD) after regressing out age and sex. **Panel B:** Increase in predictive accuracy when adding the PheRS to a baseline model with age and sex. The meta-analyzed c-indices and 95% CIs of the baseline model (x-axis) compared to a model with added PheRS (y-axis).

We found that, in FinnGen, all PheRS were correlated, mostly positively, with the total number of phecodes an individual had recorded (Persons’ r ranging from 0.77 for asthma to –0.43 for breast cancer; Supplement Figure 1C). To further test whether this meant that the PheRS are more predictive in older individuals who have had more time to accumulate diagnoses in their EHR, we stratified the FinnGen test set to a younger group aged 32-51 and an older group aged 52-70 years. However, unexpectedly, we found a significantly stronger association of the PheRS in the younger age group for 4/13 disease and only for breast cancer was the relative risk in the older group significantly larger than for the younger group, while no differences were observed in the remaining diseases (Supplement Figure 4).

### PheRS transfer well between studies

We examined PheRS transferability by comparing, in FinnGen, externally– and internally-trained PheRS. Externally-trained PheRS were trained on the training set of the UKB and EstB study and tested on the same test set as the FinnGen internally-trained PheRS. Externally-trained PheRS were moderate to strongly correlated with internally-trained PheRS (average Pearsons’ *r*=0.45, range –0.09-0.74; **Figure 3A**, Supplement Table 5). Not surprisingly, PheRS that were poor predictors of the disease were also poorly correlated between their internally-trained and externally-trained versions (i.e. colorectal and breast cancer). Most externally-trained PheRS were significantly associated with disease risk in FinnGen (**Figure 3B**) and showed significant improvements in c-index over age and sex (Supplement Figure 5). In some instances, as in the case of the PheRS models for MDD and CHD trained in the UKB and the gout models trained in the UKB and EstB, externally-trained PheRS c-index improvements were not significantly different from those achieved by the FinnGen internally-trained PheRS. Nonetheless, we observed that most PheRS disease associations were significantly lower with the externally-trained PheRS (**Figure 3B**).

**Figure 3:**
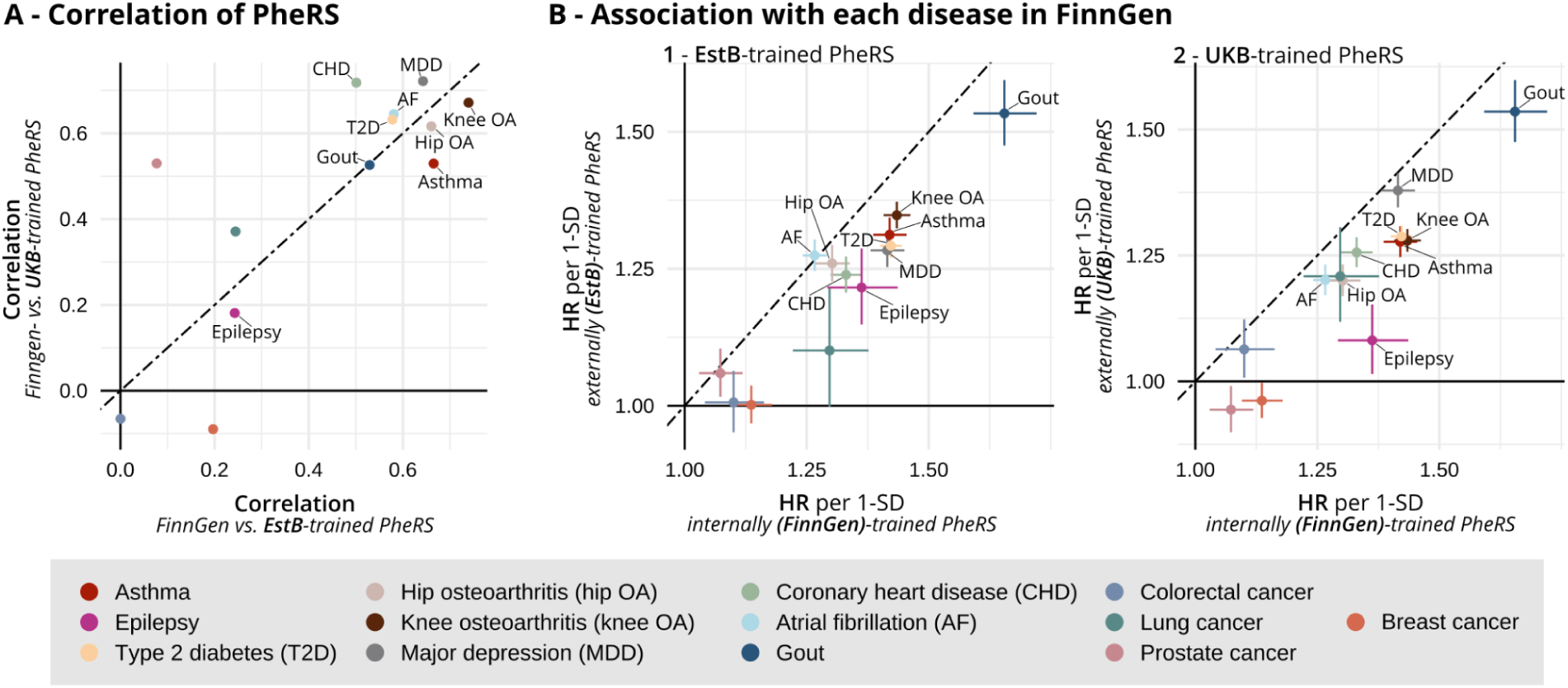
PheRS validation in FinnGen. **Panel A**: Correlation (Persons’ *r*) between the internally-trained PheRS in FinnGen and the externally-trained PheRS, tested in FinnGen. Externally-trained PheRS were trained in 50% of individuals in UKB (y-axis) and EstB (x-axis). **Panel B:** Association of FinnGen internally-trained PheRS with each disease compared to the externally-trained models. HRs and 95% CIs of the FinnGen-trained PheRS (x-axis) vs. the externally-trained PheRS (y-axis), with EstB on the left and UKB on the right. The HRs are shown for a 1-SD increase of the PheRS after regressing out age and sex.

### Phecode importance varies across studies

Despite good PheRS transferability, we found marked differences in the prevalence of different phecodes between the studies (Supplement Table 6). When considering codes with a prevalence of at least 1%, only 20% of phecodes (N=49) could be observed in all three studies (**Figure 4A**). These differences can be partially explained by different types of diagnostic information from the EHR available in each study. For example, the inclusion of primary care diagnoses in the EstB study leads to a higher number of phecodes, with 32% (N=77) unique to that study (**Figure 4A+B**). The FinnGen and UKB studies, on the other hand, only utilized diagnoses from secondary care.

**Figure 4:**
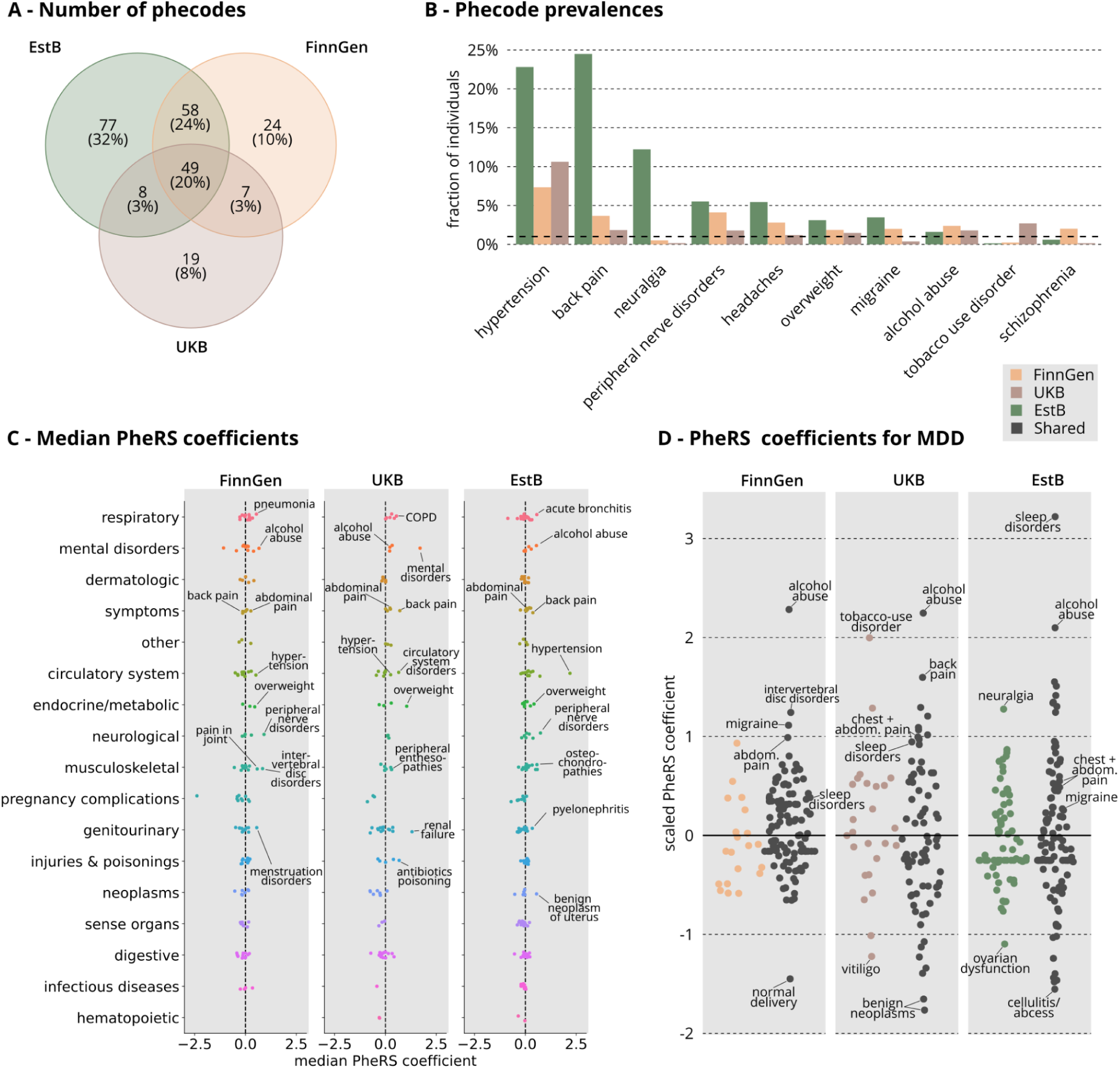
Phecode prevalence and coefficients in each study. **Panel A**: A Venn diagram showing the number of phecodes present in each of the three studies and shared between all combinations of the three studies. We only considered phecodes with a prevalence >1% in each study. Yellow color indicates FinnGen-specific codes, brown UKB-specific codes, green EstB-specific codes and black codes present in all three studies. The same color coding applies to panels B and D. **Panel B:** Phecode prevalences for selected example codes in the three studies. The black dashed line indicates a prevalence of 1%. **Panel C:** Median of PheRS coefficients over the 13 diseases in each study. Only coefficients used by at least 7/13 models in the studies are shown (see Methods for phecode exclusion rules in the PheRS models). Different colors and the y-axis labels indicate different phecode categories. Black dashed lines correspond to coefficient values of 0. **Panel D:** A detailed look at all the PheRS coefficients for major depression (MDD) in the three studies. Black color marks common phecodes in the MDD PheRS models across the studies, while other colors indicate biobank-specific codes (yellow=FinnGen, brown=UKB, green=EstB). PheRS coefficients are standardized to 0 mean and 1 standard deviation for each model separately for easier comparison of coefficient importance across the studies.

The set of phecodes unique to each study included important predictors for many of the diseases. To highlight one example in each study, we found neuralgia (code 766) to be among the top 20 predictors for hip OA, CHD, and MDD in the EstB. In FinnGen, schizophrenia (code 295) was an important predictor in T2D, lung cancer, and epilepsy; and in the UKB, tobacco use disorder (code 318) was among the most important predictors for hip OA, T2D, lung cancer, CHD, and MDD. However, other predictors such as hypertension (code 401), overweight (code 278), alcohol abuse (code 317), and peripheral nerve disorders (code 351) were prevalent diagnoses in all three studies and showed a large consistent effect across diseases (**Figure 4B+C**, Supplement Table 7).

We took a closer look at the top predictors in the individual PheRS models. **Figure 4D** shows the shared and study-specific predictors in the PheRS models for major depression (MDD). We found that the top predictors in each PheRS captured three main categories: substance abuse, sleep disorders and pain-related problems. The most consistent phecode related to substance abuse in all three studies was *alcohol abuse* (code 317; FinnGen rank 3, UKB rank 2, and EstB rank 4), while other diagnoses such as *tobacco use disorder* (code 318) were only captured in the UKB study (rank 3). The most important predictors related to pain disorders in FinnGen were *intervertebral disc disorders* (code 722, rank 4) and *migraine* (code 340, rank 5), while in the UKB it was *back pain* (code 760, rank 4) and in the EstB *peripheral nerve disorders* (code 351, rank 9) and *other headache syndromes* (code 229, rank 10). Nevertheless, while the list of most important predictors varied, each of the PheRS models also captured other pain-related diagnoses with lower ranks. Supplement Figure 6 shows, for 6 additional diseases, how common and study-specific phecodes contribute to PheRS prediction.

### PGS and PheRS are orthogonal predictors

Finally, we compared the PheRS and corresponding PGS associations. Both were significantly associated with all diseases in the meta-analysis (p<0.05). However, the magnitude of the associations varied across diseases. For 4 out of the 13 diseases (epilepsy, MDD, knee OA, and lung cancer), the PheRS showed a stronger association with the diseases than the PGS, and for 4/13 there was no significant difference (Supplement Figure 7C). However, when looking at the top 10% of most at-risk individuals compared to the 20% at average risk, the PheRS capture the risk better for 8/13 diseases (T2D, gout, lung cancer, asthma, MDD, epilepsy, hip OA, and knee OA; **Figure 5A**, Supplement Figure 6A). Moreover, PheRS provided additional information on top of PGS. Adding PGS to a model with PheRS, age, and sex led to significant improvements for 10/13 in FinnGen, 3/4 in the UKB, and 6/13 in the EstB (**Supplementary** Figure 7A). Similarly, adding the PheRS to a model with PGS, age, and sex significantly increased the c-index for 9/13 diseases in FinnGen, 2/4 diseases in the UKB, and 6/13 diseases in the EstB (**Supplementary** Figure 7B). The number of diseases with significant improvements due to adding the PheRS is similar to that achieved when adding the PheRS to age and sex (**Supplement Figure 2A, see Methods and Supplementary Text for more details**)

**Figure 5:**
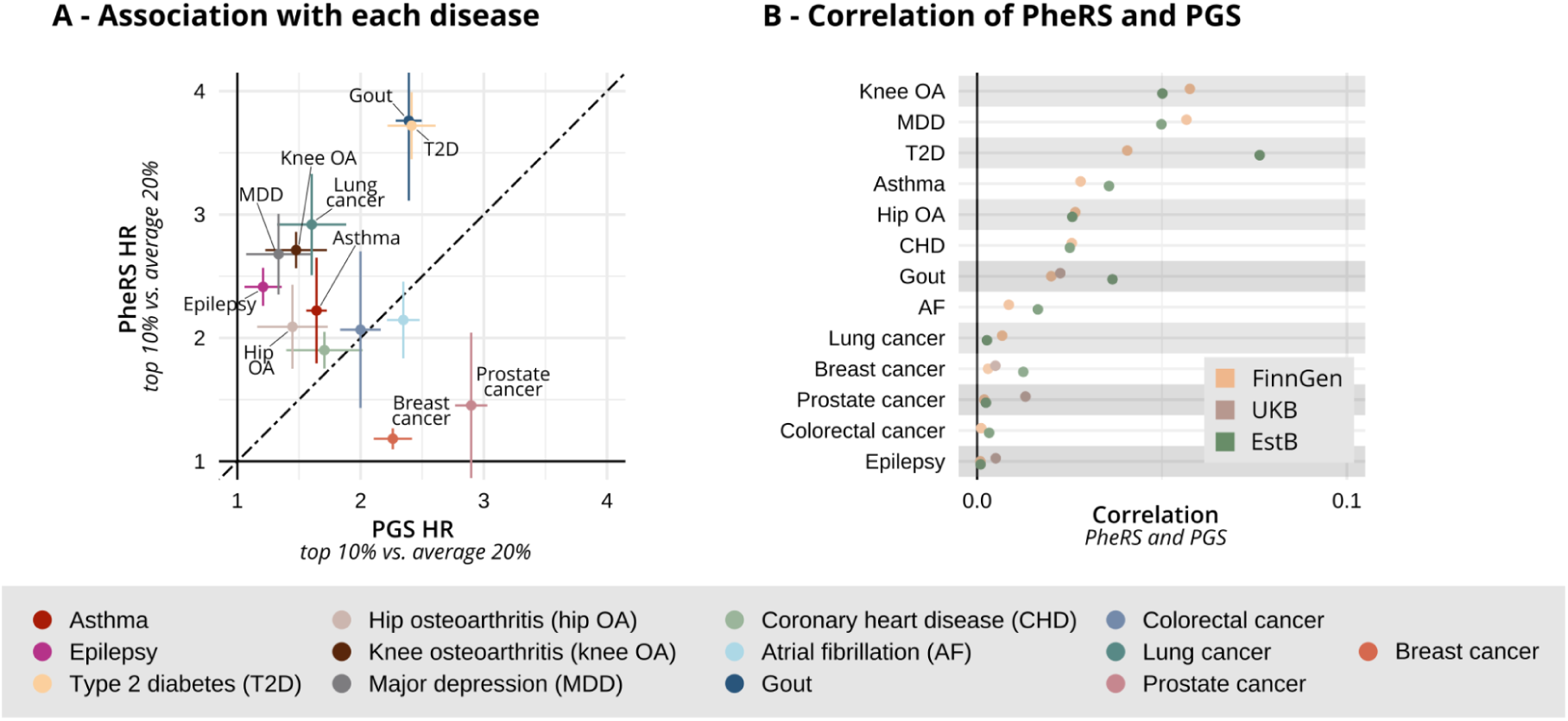
Comparison of PGS and PheRS. **Panel A**: Association of PGS (x-axis) and PheRS (y-axis) scores with each disease. The meta-analyzed HRs (95% CI) for the top 10% at risk compared to the average 20% based on the scores after regression out age, sex, and the first 10 PCs. **Panel B:** Correlation (Persons’ *r*) between the PheRS and PGS scores separately in each study (FinnGen: yellow, UKB: brown, EstB: green). Due to sample overlap with the GWASs, PGS could only be calculated for 4 diseases in UKB (see Methods for details).

Overall, we found that the EHR data and genetic information capture largely orthogonal information as shown by the low correlation between the two scores (average Pearsons’ *r*=0.02, range 0.00-0.08, **Figure 5B**).

## Discussion

In this study, we investigated the accuracy and transferability of EHR-based models (PheRS) in predicting the 8-year risk for 13 common diseases in three large biobank-based studies (FinnGen, EstB, and UKB) compared to PGS. Our results highlight the complementarity of PheRS and PGS for a range of diseases, suggesting that combining EHR and genetic data can be an advantageous strategy for the prediction of many common diseases. Both PheRS and PGS were derived to be independent from age and sex effects, thus providing orthogonal information to these two key risk factors. Furthermore, we were able to successfully validate in FinnGen the models trained in EstB and UKB, suggesting that the PheRS models capture relevant risk factors that are not only study– or healthcare system-specific.

While the performance of the PheRS models varied between diseases, the PheRS for asthma, MDD, T2D, knee OA, and gout, in particular, performed well across all three studies. In contrast, colorectal cancer, prostate cancer, and breast cancer PheRS models performed poorly, likely also due to the low case counts in our data. We expected to see low transferability of the PheRS between studies due to differences in clinical and disease coding practices in different countries and healthcare systems. Nonetheless, we found that the PheRS replicated well for many of the diseases although, as expected, most PheRS trained within-study performed better. The good transferability of PheRS was also surprising given the large variability in prevalence of phecodes we found across studies, with only 20% of them observed in all three studies. However, our results are in line with a few previous studies that show that it is possible to create predictors that are generalizable across healthcare systems^6–8,31^.

Looking closer at the phecodes prevalent in each study and their importance in the PheRS models, we find that the PheRS models that transfer well from the UKB or EstB to FinnGen utilize both study-specific phecodes, and phecodes shared between the studies. In some cases, such as gout, a large part of the transferability of the models could already be explained by a few major risk factors such as hypertension, high BMI, and diabetes that are consistent in all three studies^50^. In other cases, the relevance of each predictor was more intricate. For example, in UKB, one of the most important phecodes for major depression (MDD) was *tobacco use disorder*, but this code had very low prevalence in FinnGen. Instead, we found that the phecodes for *alcohol abuse* (code 317) was a prevalent and important predictor in all three studies. Both alcohol abuse and sleep disorders, another important and prevalent predictor in all the studies, are known complex comorbidities of MDD^51^. We hypothesize that many of the different phecodes captured a single underlying risk factor. For example, several different pain-related diagnoses were among the top predictors for MDD, each likely capturing underlying pain problems^52^, with the top predictors differing between models. The elastic net penalty allows a non-zero coefficient for many correlating phecodes, which alleviates the issue of the same underlying medical issue being coded differently in different EHRs. This suggests that leveraging similarity between diagnostic codes is an important aspect in creating transferable EHR-based predictors.

We kept the PheRS approach simple to demonstrate its feasibility. More complex models could further exploit the longitudinal nature of EHR information and utilize other data modalities available in the EHR-systems^5,30,53^. Further, by using a 2-year washout period and excluding very closely related conditions from the predictors, we remained conservative in removing co-morbidities directly related to the disease. Without this buffer the performance of the models will likely increase and be more relevant in a clinical context^6^. To improve generalizability of the models we collapsed phecodes into the first three digits to reduce the effect of different diagnostic codes being used in different countries to describe the same underlying phenomenon^54^. For example, in the EstB study phecodes *hypertensive heart disease* (code 401.21) and *essential hypertension* (code 401.1) were equally prevalent diagnoses capturing the risk factor hypertension (code 401), while in FinnGen *hypertensive heart disease* (code 401.21) had a prevalence of <1%. Other approaches could include mapping diagnostic codes to OMOP-concepts, which has been shown to facilitate EHR-based models that transfer between different countries^31^.

Importantly, while we did not exclude individuals based on their genetic ancestry, the UKB still consists mainly, and FinnGen and the EstB almost exclusively, of individuals of European ancestry. Thus, our study does not properly assess the important issue that individuals of different ethnicities face inequalities in healthcare access^55,56^. Important open questions for future work, in addition to the generalizability of EHR-based scores for non-European genetic ancestries, include how to optimally model diagnostic codes for best generalizability as well as leveraging data from different and diverse cohorts with for example federated learning approaches^57^.

To our knowledge, correlation between PGS and EHR-based scores have not been comprehensively studied. For CAD, Petrazzini et al. ^7^ found that the inclusion of PGS did not improve prediction compared to an EHR-based score, while Zhao et al. ^6^ found that the inclusion of genetic information significantly improved models with both EHR-based predictors and the gold standard model for CAD risk prediction (ACC/AHA) Pooled Cohort Risk Equations). For 8/13 diseases studied here, we observe a significant improvement in onset prediction when integrating PheRS on top of PGS. While for many of the cancers (colorectal cancer, prostate cancer, and breast cancer), the PGS were more informative, for diseases such as MDD, epilepsy, and knee OA the PheRS better captured the risk. Interestingly, PheRS were specifically better than PGS in capturing high-risk individuals. Individuals in the top 10% of PheRS had higher HR than those in the top 10% of PGS for 8/13 disease, probably reflecting those individuals with key co-morbidities. Further, we observe very low correlation between PGS and PheRS, indicating that these two data sources contain largely independent information that is predictive of disease onset. A few prior studies on the interaction between PGS and selected risk factors found no evidence for interaction^11,51^.

Patient’s diagnostic history has always been a key piece of information for medical professionals when considering future treatment. As we are moving towards translating PGS to clinical use, it is worth considering integrating, in a comprehensive manner, also the information about an individual’s diagnosis history which in many countries is already collected in a centralized electronic manner. This would not be a large shift from current practice, as selected comorbidities are used in many clinical risk stratification algorithms, for example, QRisk^58^ for evaluating risk of heart attack or stroke in 10 years, or QDiabetes for evaluating 10 year risk of T2D^59^. A recent study (Steinfeld et al. ^60^), showed that EHR-based models trained specifically to predict risk of five different cardiovascular events performed similarly or better than conventional risk scores (QRISK3, ASCVD and SCORE2)^31^. Similarly, Zhao et al.^6^ found that the machine learning models trained on longitudinal EHR data outperformed the gold standard risk model (ACC/AHA) Pooled Cohort Risk Equations) for the prediction of cardiovascular disease. These comparisons are interesting for diseases with established risk scores. However, for many of the diseases studied here there are no established risk algorithms, making an EHR-based risk stratification approach even more relevant.

In this study we show that, across many diseases and multiple studies with different underlying healthcare systems and EHRs, relatively simple elastic net-based risk scores that consider an individual’s previous diagnosis history can improve disease risk prediction when combined with PGS. Information already available from the EHR provides orthogonal information to PGS and could be a cost-effective approach for risk estimation.

## Methods

### Study setup

As outlined in **Figure 1B**, each study consisted of a 10-year observation (6-year for EstBB due to shorter follow-up) and an 8-year prediction period, separated by a 2-year washout period. Each disease’s case and control definitions were based on diagnoses acquired in the 8-year prediction period (2011/01/01 –> 2019/01/01). The ICD-codes used to define the cases for each disease were based on previous harmonization between FinnGen and the EstBB phenotypes by the INTERVENE consortium^46^ (Supplement Table 9). We consider all individuals as controls that are not cases. We only considered adults aged 32-70 in 2011/01/01 and removed all individuals diagnosed with the disease before this time. The lower limit for age of inclusion was chosen due to the inclusion of education level in some of the models and determined based on the median age of obtaining a doctoral degree in the FinnGen dataset. Using this lower limit, most individuals included have finished their highest level of education. Further, we remove all individuals with a diagnosis outside the prediction period (2011/01/01 –> 2019/01/01) and those lost to follow-up before the start of the prediction period. The ICD-codes used to define the cases for each disease and the number of cases and controls in each study are listed in Supplement Tables 9+2.

We included 845,929 individuals (Supplement Table 1) from 3 biobank-based studies: FinnGen^41^, UKB^40^ and EstB ^34^ linked with national registers or EHRs. In FinnGen we used Data Freeze 10, which includes 412,090 individuals, of which 266,179 were aged 32-70 in 2011/01/01. The longitudinal ICD-code diagnoses used to define the phecodes and the case and control status for each disease were based on in– and outpatient hospital register information. The UKB study included 464,076 individuals aged 40-70, with the ICD-code diagnoses based on inpatient information. The EstB study included 199,868 individuals of which 115,674 were aged 32-70. Here we also had primary care data as well as self-reported diagnoses available. More details on the phenotype harmonization can be found from Jermy *et al.*^46^ and the Supplement Methods.

### Predictors

#### PGS

The PGS were previously computed by the INTERVENE consortium^46^ and based on the recent publicly available genome-wide association study (GWAS) summary statistics, with minimal overlap with our study cohorts (Supplement Table 10) using MegaPRS ^61^ with the BLD-LDAK heritability model. For the Cox-PH models we removed individuals from the studies that were part of the GWAS on which the PGS were based. Due to the large overlap with the UKB individuals, we only had PGS for gout, epilepsy, breast and prostate cancer available in the UKB.

#### PheRS

For the EHR-based models, we trained elastic net models^43^ on ICD-9 and ICD-10 diagnoses mapped to phecodes. The phecode mapping was based on the v1.2b1 of the phecode map^44,47^ from https://phewascatalog.org/, with some manual additions. Since we only considered diagnoses during the observation period starting in 1999, all diagnoses were ICD-10 based in our data. To get the most complete mapping we removed all special characters from the ICD-code and then if we could not find a match in the phecode map, we shortened the code by one digit until it could either be mapped or had to be removed. The complete mapping used can be found from Supplement Table 11. As our target phenotypes were defined based on ICD-codes we exclude predictors part of the exclusion range of the phecodes separately for each phenotype (Supplement Table 12). We only considered phecodes with a prevalence at least 1% of the study population (Supplement Table 6).

We implemented the PheRS using the LogisticRegression function from scikit-learn (version 1.3.2)^62^. We included age (at the start of the prediction period 2011/01/01) and sex as predictors in the PheRS models because they are important predictors and otherwise the models would reconstruct predictors for age and sex using combinations of the phecode diagnoses, which would make interpretation of the phecode coefficient values challenging. Nonetheless, age and sex effect were then regressed out when evaluating the performances of the PheRS (see below). Models were penalized with the elastic net penalty. Predictors were coded as 1/0, where 1=”predictor observed during the observation window” and 0=”predictor not observed during the observation window”, for each disease separately. For training, 50% of the data was used, and this was further divided into training (85%) and hold-out test (15%) sets. Sizes of the training data sets are shown for each disease and study Supplement Table 2. L1 to L2 ratio hyperparameter of the elastic net models was optimized using grid search and 5-fold cross-validation over the range 0.05-0.95 (step size = 0.05), simultaneously with inverse of the regularization strength (C) over possible values: 1e-5, 5e-5, 1e-4, 5e-4, 1e-3, 5e-3, 1e-2, 5e-2, 1e-1, 5e-1, 1. Balanced class weights were used, based on class frequencies in the training data.

Model fitting was done using stochastic average gradient descent. Best L1 to L2 ratio was selected based on the average precision score using 5-fold cross validation on the training split. Missing values of predictors were imputed to the mean of the corresponding predictor in the study-specific training data and all predictors were standardized to zero mean and unit variance on the study-specific training data prior to model fitting. The code for training the PheRS models is available at: https://github.com/intervene-EU-H2020/INTERVENE_PheRS.

The PheRS models trained within the UKB or the EstBB data on 50% of individuals were used to make predictions in FinnGen test set as is without any retraining with FinnGen data. Standardization and imputation were performed based on the biobank-specific training data, meaning that e.g. when assessing the performance of the UKB-trained model in FinnGen, the FinnGen test set data was imputed and standardized based on the feature-specific means and standard deviations from the UKB.

### Cox proportional-hazards models

Ultimately, each individual was assigned 13 different PGS and PheRS scores describing their risk of getting a disease diagnosis in the prediction period based on genetic or EHR-based information. To make the PheRS and PGS comparable we regressed out the effect of age, sex and the first 10 genetic PCs from all continuous scores using the residuals from a logistic regression with the score as outcome. Subsequently we scaled all predictors to have a mean of zero and standard deviation of 1. We then used these scores in separate Cox proportional-hazards models (Cox-PH), with the survival time defined as the time from 2011 until either diagnosis, censoring (end of follow-up), or the end of the prediction period.

Additionally, we considered the Charlson-Comorbidity Index (CCI)^48,49^ – developed to account for the individual’s overall comorbidity burden – and individuals highest achieved education level in 2011 – an indicator of their socio-economic status. For the CCI we compared the top 10% of individuals with the highest CCI to the rest. The high-risk group included individuals with a CCI>=2 and a few younger ones with a CCI of 1. For the highest education level we mapped each study’s education coding to the 2011 International Standard Classification of Education (ISCED-11; Supplement Table 13) codes. We compared the risk of individuals with basic education (ISCED-11: 1-4) to those who achieved high education levels (ISCED-11: 5-7).

We used the survival^63^ package in R for creating the Cox-PH models and the Hmisc^64^ package to calculate the c-indices and 95% CIs. For a Cox-PH model with binary outcomes, the predicted survival times can be shown to be equal to the survival probability, so the c-index is equivalent to the area under the curve of the receiver operating characteristic curve (AUCROC)^65,66^. The meta-analysis of the HRs and c-indices was performed using the metafor^67,68^ package in R with a random effects model. We used two-tailed p-values based on the z-scores to compare the difference in HR magnitude and significant increases in the c-index.

### Comparison of phecode coefficients between different PheRS models

The elastic net hyperparameters were separately optimized for each PheRS model. This means that the absolute magnitudes of the coefficients for phecodes are not comparable between different PheRS. However, the relative importances of phecodes can still be compared, i.e. whether for example the same phecodes are among the most important predictors in two different PheRS. To make visualization of the phecode importances in different PheRS clearer, we standardized the coefficients of each PheRS separately to a mean of 0 and a standard deviation of 1 for the display items. Further, in each study we ranked the phecodes in descending order by the PheRS coefficient values and assigned them ascending ranks. Thus, a lower rank indicates a higher PheRS coefficient in the model. Both the unscaled PheRS coefficients and ranks are Supplement Table 7.

## Author contributions

KED and TH contributed equally. KED, TH and AG wrote the manuscript with significant input from all the other authors. TH and KED wrote the PheRS code, KED wrote the Cox model code, BJ contributed the PGS. KED and TH performed the analyses in FinnGen, TH, KED in the UK Biobank and MTL and KL in the Estonian Biobank. KED combined the results and created the figures together with TH. KED, TH and ZY preprocessed the FinnGen and the UK Biobank data, MTL and KL preprocessed the Estonian Biobank data. TH, BJ and AG developed the original idea. AG, SR and RM supervised the study.

## Supporting information

Supplement Methods and Results

## Data Availability

The individual-level data in these studies is protected for data privacy, access is regulated through the biobanks. The Finnish biobank data can be accessed through the Fingenious services (https://site.fingenious.fi/en/) managed by FINBB. Researchers interested in EstBB can request access at https://www.geenivaramu.ee/en/access-biobank and the UKB data are available through a procedure described at http://www.ukbiobank.ac. The GWAS data used in this study are available in the GWAS catalog database under accession codes listed in Supplement Table 10. The PGS scores generated in this study are available in the PGS Catalog under publication ID: PGP000618 and score IDs: PGS004869-PGS004886.

https://github.com/intervene-EU-H2020/INTERVENE_PheRS

https://github.com/intervene-EU-H2020/onset_prediction

## Acknowledgements

We want to acknowledge the participants and investigators of the FinnGen, UK Biobank and the Estonian Biobank studies.

This study has received funding from the European Union’s Horizon 2020 research and innovation programme under grant agreement no. 101016775. This Estonian Biobank study was funded by the European Union through the European Regional Development Fund Project No. 2014-2020.4.01.15-0012 GENTRANSMED. A.G. has received funding from the European Union’s Horizon 2020 research and innovation programme under grant no. 101016775, the European Research Council under the European Union’s Horizon 2020 research and innovation programme (grant number 945733) and from Academy of Finland fellowship grant no. 323116.

We want to acknowledge the participants and investigators of the FinnGen study listed in Supplement Figure 14. The FinnGen project is funded by two grants from Business Finland (HUS 4685/31/2016 and UH 4386/31/2016) and the following industry partners: AbbVie Inc., AstraZeneca UK Ltd, Biogen MA Inc., Bristol Myers Squibb (and Celgene Corporation & Celgene International II Sàrl), Genentech Inc., Merck Sharp & Dohme LCC, Pfizer Inc., GlaxoSmithKline Intellectual Property Development Ltd., Sanofi US Services Inc., Maze Therapeutics Inc., Janssen Biotech Inc, Novartis AG, and Boehringer Ingelheim International GmbH. Following biobanks are acknowledged for delivering biobank samples to FinnGen: Auria Biobank (www.auria.fi/biopankki), THL Biobank (www.thl.fi/biobank), Helsinki Biobank (www.helsinginbiopankki.fi), Biobank Borealis of Northern Finland (https://www.ppshp.fi/Tutkimus-ja-opetus/Biopankki/Pages/Biobank-Borealis-briefly-in-English.aspx), Finnish Clinical Biobank Tampere (www.tays.fi/en-US/Research_and_development/Finnish_Clinical_Biobank_Tampere), Biobank of Eastern Finland (www.ita-suomenbiopankki.fi/en), Central Finland Biobank (www.ksshp.fi/fi-FI/Potilaalle/Biopankki), Finnish Red Cross Blood Service Biobank (www.veripalvelu.fi/verenluovutus/biopankkitoiminta), Terveystalo Biobank (www.terveystalo.com/fi/Yritystietoa/Terveystalo-Biopankki/Biopankki/) and Arctic Biobank (https://www.oulu.fi/en/university/faculties-and-units/faculty-medicine/northern-finland-birth-cohorts-and-arctic-biobank). All Finnish Biobanks are members of BBMRI.fi infrastructure (https://www.bbmri-eric.eu/national-nodes/finland/). Finnish Biobank Cooperative –FINBB (https://finbb.fi/) is the coordinator of BBMRI-ERIC operations in Finland.

We thank participants and scientists involved in making the UK Biobank resource available (http://www.ukbiobank.ac.uk/). This research has been conducted using the UKB resource under approved application number 78537.

The EstBB research team received funding from the Estonian Research Council grant TT17 “Estonian Centre for Genomics”. Data analysis was carried out in part in the High-Performance Computing Center of the University of Tartu. K.L. and R.M. received funding from the Estonian Research Council grant PUT (PRG1911) and the Estonian Research Council grant TK (TK214).

We acknowledge CSC—IT Center for Science, Finland, for computational resources.

The funders had no role in study design, data collection and analysis, decision to publish or preparation of the manuscript.

## Data and code availability

The code for PheRS model training is available at https://github.com/intervene-EU-H2020/INTERVENE_PheRS and for the Cox-PH models as well as the final analysis of results at https://github.com/intervene-EU-H2020/onset_prediction.

The individual-level data in these studies is protected for data privacy, access is regulated through the biobanks. The Finnish biobank data can be accessed through the Fingenious^®^ services (https://site.fingenious.fi/en/) managed by FINBB. Researchers interested in EstBB can request access at https://www.geenivaramu.ee/en/access-biobank and the UKB data are available through a procedure described at http://www.ukbiobank.ac. The GWAS data used in this study are available in the GWAS catalog database under accession codes listed in Supplement Table 10. The PGS scores generated in this study are available in the PGS Catalog under publication ID: PGP000618 and score IDs: PGS004869-PGS004886.

## Ethics declarations

Patients and control subjects in FinnGen provided informed consent for biobank research, based on the Finnish Biobank Act. Alternatively, separate research cohorts, collected prior to the Finnish Biobank Act came into effect (in September 2013) and the start of FinnGen (August 2017), were collected based on study-specific consents and later transferred to the Finnish Biobanks after approval by Fimea (Finnish Medicines Agency), the National Supervisory Authority for Welfare and Health. Recruitment protocols followed the biobank protocols approved by Fimea. The Coordinating Ethics Committee of the Hospital District of Helsinki and Uusimaa (HUS) statement number for the FinnGen study is Nr HUS/990/2017. The FinnGen study is approved by Finnish Institute for Health and Welfare (permit numbers: THL/2031/6.02.00/2017, THL/1101/5.05.00/2017, THL/341/6.02.00/2018, THL/2222/6.02.00/2018, THL/283/6.02.00/2019, THL/1721/5.05.00/2019 and THL/1524/5.05.00/2020), digital and population data service agency (permit numbers: VRK43431/2017-3, VRK/6909/2018-3, VRK/4415/2019-3), the Social Insurance Institution (permit numbers: KELA 58/522/2017, KELA 131/522/2018, KELA 70/522/2019, KELA 98/522/2019, KELA 134/522/2019, KELA 138/522/2019, KELA 2/522/2020, KELA 16/522/2020), Findata permit numbers THL/2364/14.02/2020, THL/4055/14.06.00/2020, THL/3433/14.06.00/2020, THL/4432/14.06/2020, THL/5189/14.06/2020, THL/5894/14.06.00/2020, THL/6619/14.06.00/2020, THL/209/14.06.00/2021, THL/688/14.06.00/2021, THL/1284/14.06.00/2021, THL/1965/14.06.00/2021, THL/5546/14.02.00/2020, THL/2658/14.06.00/2021, THL/4235/14.06.00/2021, Statistics Finland (permit numbers: TK-53-1041-17 and TK/143/07.03.00/2020 (earlier TK-53-90-20) TK/1735/07.03.00/2021, TK/3112/07.03.00/2021) and Finnish Registry for Kidney Diseases permission/extract from the meeting minutes on 4th July 2019. The Biobank Access Decisions for FinnGen samples and data utilized in FinnGen Data Freeze 10 include: THL Biobank BB2017_55, BB2017_111, BB2018_19, BB_2018_34, BB_2018_67, BB2018_71, BB2019_7, BB2019_8, BB2019_26, BB2020_1, BB2021_65, Finnish Red Cross Blood Service Biobank 7.12.2017, Helsinki Biobank HUS/359/2017, HUS/248/2020, HUS/150/2022 § 12, §13, §14, §15, §16, §17, §18, and §23, Auria Biobank AB17-5154 and amendment #1 (August 17 2020) and amendments BB_2021-0140, BB_2021-0156 (August 26 2021, Feb 2 2022), BB_2021-0169, BB_2021-0179, BB_2021-0161, AB20-5926 and amendment #1 (April 23 2020)and it’s modification (Sep 22 2021), Biobank Borealis of Northern Finland_2017_1013, 2021_5010, 2021_5018, 2021_5015, 2021_5023, 2021_5017, 2022_6001, Biobank of Eastern Finland 1186/2018 and amendment 22 § /2020, 53§/2021, 13§/2022, 14§/2022, 15§/2022, Finnish Clinical Biobank Tampere MH0004 and amendments (21.02.2020 and 06.10.2020), §8/2021, §9/2022, §10/2022, §12/2022, §20/2022, §21/2022, §22/2022, §23/2022, Central Finland Biobank 1-2017, and Terveystalo Biobank STB 2018001 and amendment 25th Aug 2020, Finnish Hematological Registry and Clinical Biobank decision 18th June 2021, Arctic Biobank P0844: ARC_2021_1001.

Ethics approval for the UK Biobank study was obtained from the North West Centre for Research Ethics Committee (11/NW/0382). UK Biobank data used in this study were obtained under approved application 78537.

The activities of the EstBB are regulated by the Human Genes Research Act, which was adopted in 2000 specifically for the operations of the EstBB. Individual level data analysis in the EstBB was carried out under ethical approval 1.1-12/624 from the Estonian Committee on Bioethics and Human Research (Estonian Ministry of Social Affairs), using data according to release application S22, document number 6-7/GI/16259 from the EstBB.

Andrea Ganna is the founder of Real World Genetics Oy. Bradley Jermy became an employee of BioMarin after his part of this work was completed. Kristi Läll has participated as an analyst in a collaboration research project at the Institute of Genomics, University of Tartu, which was funded by Geneto OÜ. No other authors have competing interests to declare.

