## Supplement Methods and Results for "Transferability and accuracy of electronic health record-based predictors compared to polygenic scores"

### Supplement Methods
#### Registry data
##### FinnGen
The FinnGen data was the same as was used by Jermy *et al.*[1], the following copied from the Supplement Material.
Phenotype data within FinnGen is constructed from the collection of nationwide electronic health registers. This gives a comprehensive coverage of almost all of a patient's interactions with the health service including hospitalizations, medications, procedures and deaths. The 18 different registers used by the project are listed below in order of their follow-up times:

- Finnish Cancer Registry - From 1953
- Register of Congenital Malformations - From 1963
- Reimbursement - From 1964
- Population Register - From 1964
- Finnish Registry for Kidney Diseases - From 1964
- Causes of Death - From 1969
- Care Register for Health Care Inpatient Visits, HILMO - From 1969
- Socio-economic data - From 1970
- The Finnish Registry of Visual Impairment - From 1983
- Medical Birth Register - From 1987
- Finnish National Infectious Disease Register - From 1989
- Cervical Cancer Screening - From 1991
- Breast Cancer Screening - From 1992
- Drug Purchases - From 1995
- The Care Register for Social Welfare - From 1995
- Care Register for Health Care, specialist outpatient visits, HILMO - From 1998
- Register of Primary Health Care Visits, Avohilmo - From 2011
- The Finnish Vaccination Register - From 2011

Note: while primary health care visits are included within FinnGen, by default these cases are excluded from the endpoints. As such, we only consider secondary care data for our disease endpoints.

##### UK Biobank
All data is based on hospital records. We considered all diagnoses marked as primary, secondary and tertiary and took the earliest date of diagnoses from any of the columns.
Data columns used:

- HESIN the hospital inpatient admission with the main table (field 1063) containing information on all hospital inpatient admissions and the diagnosis table (field 1067) covering all diagnosis codes recorded in an inpatient admission. The date was based on the start of the hospital stay. (See also https://biobank.ndph.ox.ac.uk/ukb/refer.cgi?id=593)
- Summary ICD-10 main diagnoses (field 41202 with date 41262)
- Summary ICD-9 main diagnoses (field 41203 with date 41263)
- Summary ICD-10 secondary diagnoses (field 41204 with date 41280)
- Summary ICD-9 secondary diagnoses (field 41205 with date 41281)

- Cause of death (field 40001 for main ICD-10 and 40002 for secondary ICD-10 with dates from 40000)

Additional variables were extracted from:

- Date of birth: based on month of birth (field 52), year of birth (field 34) and then the 15th of each month.
- PCs: Genetic principal components from (field 22009)
- End of followup: Based on information from date of death (field 40000)
- Education: Qualifications (field 6138) mapped to ISCED-11 with this mapping: Supplement Table 13

All elements were constructed using information from 0 - initial assessment visit (2006-2010) at which participants were recruited and consent given, if 0 not available then 1 - first repeat assessment visit (2012-13) and otherwise 2 - imaging visit (2014+) if neither 0 or 1 contained information.

For the code and further documentation see: GitHub - UKB phenotyping.

**EstB**

The EstB data was the same as was used by Jermy *et al.*[1], the following copied from the Supplement Material.

Phenotype data within EstBB is put together from the collection of electronic health registers, including from two largest hospitals in Estonia. We include both primary and secondary care data as well as self-reported diagnoses. Estonia has a solidary health insurance system and national public health insurance covers ~94% of the population (https://eurohealthobservatory.who.int/countries/estonia). Causes of death and Cancer registry record all cases despite the health insurance status in Estonia. Following registries were included in phenotype definition process:

- Causes of Death Registry- diagnoses from 2003 until 2020
- National Cancer Registry- diagnoses from 1955 until 2017
- Estonian Health Insurance Fund - From 2001 until 2020
- The North Estonia Medical Centre from 1993 until 2017
- Tartu University hospital from 2006 until 2017
- E-Health system from 1998 to 2020

Self-reported diagnoses' dates ranged from 1920 - 2018.

#### Supplement Results

**Supplement Table 1.** Number of individuals aged 32-70 in each study.

**Supplement Table 2.** Number of cases and controls for each disease and study.

**Supplement Table 3.** HRs from the Cox-proportional hazards models.

**Supplement Table 4.** C-index of the Cox-proportional hazards models.

**Supplement Table 5.** Correlation (Pearsons' r) of the internally (FinnGen)- and externally-trained PheRS predictions, the PheRS and PGS, the PheRS and unique number of phecodes recorded during the observation period (1/1/1999 - 31/12/2008), and the PheRS and CCI.

**Supplement Table 6.** Phecode prevalences in each study during the observation period (1/1/1999 - 31/12/2008) NAs indicate that less than 5 individuals have a diagnosis.

**Supplement Table 7.** PheRS coefficients in each study. Phecodes with a prevalence <1% in a study have NAs. For the rank, the coefficients are sorted in descending order, with the largest coefficient in a model thus having a rank of 1.

**Supplement Table 8.** Median PheRS coefficients across the three biobank studies. Only those phecodes (phenotypes) that were included in at least 7/13 PheRS models in a biobank study are included.

**Supplement Table 9.** Harmonized definitions of flagship diseases using ICD-10 and ICD-9 codes.

**Supplement Table 10.** Genome-wide association study summary statistics used to compute PGS and sample overlap with each biobank.

**Supplement Table 11.** Mapping of ICD-10 codes to phecodes.

**Supplement Table 12.** Excluded phecodes from the predictors for each disease.

**Supplement Table 13.** Mapping of study specific education codes to ISCED-2011

**Supplement Table 14.** FinnGen authors.

**Supplement Figure 1**. In FinnGen, we found c-index improvements for 9/13 diseases when adding the PheRS to a baseline model with age and sex (asthma, epilepsy, knee OA, T2D, MDD, hip OA, CHD, AF, and gout); in the UKB for 11/13 diseases (asthma, breast cancer, epilepsy, MDD, knee OA, hip OA, T2D, CHD, lung cancer, AF, and Gout), and in the EstB only for 7/13 (MDD, asthma, knee OA, hip OA, T2D, gout, AF; **Supplement Figure A-1**). When adding the PheRS to a baseline model including, additionally highest achieved education level and the Charlson comorbidity index the improvement persisted for 8/13 diseases in FinnGen (for epilepsy the increase was not significant anymore), 8/13 in the UKB (no epilepsy, gout and hip OA), and 6/13 in the EstB (no significant improvement for AF anymore; **Supplement Figure A-2**). When considering the c-index of the model with only PheRS as predictors, the results are very similar to the HRs of the PheRS (**Figure 2A**). We find the highest predictive discrimination for for gout (c-index=0.65; 95% CI: 0.63-0.68), lung cancer (c-index=0.62; 95% CI: 0.58-0.67), and MDD (c-index=0.61; 95% CI: 0.59-0.64). It is interesting to note that lung cancer is the only disease we consider where even with high HRs and good predictive discrimination, the PheRS only significantly improve prediction over age and sex in the UKB study (**Supplement Figure 1A**).

**Supplement Figure 2**. Overall, integrating the CCI only lead to minor improvements in the models' discriminative ability, with minor significant improvements for epilepsy and gout in FinnGen (2/13), MMD, T2D, and lung cancer in the UKB (3/13) and MDD in the EstB (1/13; **Supplement Figure 2A**). Additionally, we found that the PheRS captured similar information as the CCI. The PheRS and CCI were moderately correlated (average Pearsons' $r$=0.22, range 0.02-0.41, **Supplement Figure 2D**) and adding the PheRS to a model with the CCI attenuated many of the associations of the CCI with each disease (**Supplement Figure 2B**). We found that being in the top 10% of the CCI distribution, after regressing out age and sex, led to the largest HRs for lung cancer (meta-analyzed HR=2.17, 95% CI: 1.74-2.61), gout (meta-analyzed HR=2.12, 95% CI: 1.94-2.31), and epilepsy (meta-analyzed HR=1.94, 95% CI: 1.84-2.05) when compared to the rest of the population (**Supplement Figure 2B**). The top 10% corresponds largely to individuals with a CCI>=2 and a few younger ones with a CCI of 1.

**Supplement Figure 3.** Similarly, integrating education level only lead to minor improvements in the model's discriminative ability, with minor significant improvements for asthma and T2D in FinnGen (2/13), MDD and CHD in the UKB (2/13) and no improvements in the EstB (0/13; **Supplement Figure 3A**). We found that a lower education level (ISCED-11<5) was only significantly associated with 5 out of the 13 diseases in all three studies, with the highest relative risk for lung cancer (meta-analyzed HR=1.87, 95% CI: 1.51-1.49), T2D (meta-analyzed HR=1.36, 95% CI: 1.23-1.49), knee OA (meta-analyzed HR=1.25, 95% CI: 1.18-1.32; **Supplement Figure 3B**) in all three studies. The CCI and education level combined, lead to significant improvements in c-index over age and sex for 3/13 diseases (gout, epilepsy, and MDD) in all three studies. One notable improvement is the increase in c-index for MDD in the UKB (delta c-index 0.034, p=2e-13). The PheRS, however, still capture additional information on top of these two predictors as seen in **Supplement Figures 1A&2**.

**Supplement Figure 4.** As previously reported[2], we find significantly higher HRs for the PGS in younger individuals (32-51) for 7/13 (prostate, breast cancer, T2D, AF, CHD, hip OA, and knee OA). For the other 6/13 we find no significant difference to the older age group (52-70).

**Supplement Figure 5** shows the c-index improvements of the externally-trained PheRS over age and sex in FinnGen for the 9/13 diseases where we saw an improvement with the FinnGen-trained models. With the UKB-trained models we see significant improvements for 6/9 diseases (asthma, knee OA, T2D, MDD, CHD, and gout) over baseline and for 3/6 (MDD, CHD, gout) these improvements are not significantly different to those achieved by the FinnGen-trained PheRS. With the EstB-trained models we see significant improvements for 5/9 diseases (asthma, knee OA, T2D, MDD, and gout) over baseline with only the improvements for gout not significantly different to those achieved by the FinnGen-trained PheRS.

**Supplement Figure 6.** For gout the PheRS models in all three studies capture the risk factors hypertension (code 401, FinnGen rank 3, UKB rank 4, EstB rank 3), overweight (code 278, FinnGen rank 7, UKB rank 6, EstB rank 4), and diabetes (code 250, FinnGen rank 1, UKB rank 5, EstB rank 5). Notably, in FinnGen the phecode *stillbirth* (code 634) is the fourth most important predictor after *hypertension* (code 401, rank 73 in EstB and rank 118 in the UKB) and *urticaria* (code 947, rank 7) is an important predictor in the EstB PheRS model with neither stillbirth nor urticaria having an immediately clear connection to gout. In the UKB model, *renal failure* (code 585, rank 3) is the most important predictor after age and sex and this phecode was not included in the other studies (<1% prevalence in 32-70 year olds in FinnGen and EstB).

The PheRS models for asthma captured various phecodes related to respiratory infections or inflammations with *other symptoms of respiratory system* (code 512) the most important predictor in FinnGen before age and sex (UKB rank 5, EstB rank 17). This phecode captures diagnoses such as *cough* (ICD-10 R05) and *dyspnoea* (ICD-10 R06.0). In the EstB, on the other hand, the most important predictor was *acute bronchitis and bronchiolitis* (code 483, FinnGen rank 10) which could be better captured due to the availability of diagnoses from primary health care. Further, the models captured other phecodes related to allergies, such as *allergic rhinitis* (code 476, EstB rank 3, FinnGen rank 45), *urticaria* (code 947, EstB rank 16), *atopic/contact dermatitis due to other or unspecified* (code 939, FinnGen rank 4, EstB rank 11), and *chronic sinusitis* (code 475, FinnGen rank 4, EstB rank 11). In the UKB, the phecode *other diseases of respiratory system, not elsewhere classified* (code 519, UKB rank 10) further illustrates the difference in coding between the studies, as this phecode had a prevalence of less than 1% in the other two studies and includes diagnoses such as *unspecified acute lower respiratory infection* (ICD-10 J22) which overlaps with conditions captured by the phecode for *acute bronchitis and bronchiolitis*. Similarly to the MDD PheRS models, the asthma models in each study seem to capture the underlying risk factors using multiple phecodes, overall enabling better transferability between the studies. In the UKB, however, many of the top predictors in the other models have a prevalence of less than 1%. Instead, the model includes phecodes such as *overweight* (code 278, UKB rank 3, EstB rank 14, FinnGen rank 47) and *poisoning by analgesics, antipyretics, and antirheumatics* (code 965, UKB rank 7) as top predictors, while high BMI has been previously reported as a risk factor[3] the interpretation of *poisoning by analgesics, antipyretics, and antirheumatics* in relation to asthma is not immediately clear.

For knee OA many of the most important predictors were consistent important predictors across the three studies. These include phecodes for *other peripheral nerve disorders* (code 351, rank FinnGen rank 4, UKB rank 3, EstB rank 7), *peripheral enthesopathies and allied syndromes* (code 726, rank 5 in all models), *other disorders of synovium, tendon, and bursa* (code 727, FinnGen rank 8, UKB rank 8, and EstB rank 44), *spondylosis and allied disorders* (code 721, FinnGen rank 22, UKB rank 15, EstB rank 9), *injury (NOS)* (code 1009, FinnGen rank 3, UKB rank 13, EstB rank 10), *overweight* (code 279, FinnGen rank 9, UKB and EstB rank 2), *hypertension* (code 401, FinnGen rank 26, UKB rank 4, EstB rank 3), and *varicose veins* (code 454, FinnGen rank 7, UKB and EstB rank 8). Only in the FinnGen, the phecodes *pain in limb* (code 773, rank 12) and *pain in joint* (code 745, FinnGen rank 6, EstB rank 47) were among the top predictors. While *osteochondropathies* (code 732) had rank 11 in the EstB models and was not prevalent in the other two studies.

**Supplement Figure 7.** Both PGS and PheRS add information on top of the other predictor, age, and sex. Due to sample overlap with the GWASs, PGS could only be calculated for 4 diseases in UKB (see Methods for details). Adding PGS to a model with PheRS, age, and sex led to significant improvements for 10/13 in FinnGen, 3/4 in the UKB, and 6/13 in the EstB. In the meta-analysis only the improvements for breast cancer and asthma are significant. This can be explained by the difference in c-index for the baseline models with age and sex. Similarly, adding the PheRS to a model with PGS, age, and sex significantly increased the c-index for 9/13 diseases in FinnGen, 2/4 diseases in the UKB, and 6/13 diseases in the EstB. In the meta-analysis the increase for asthma and epilepsy was significant. The number of diseases with significant improvements by the PheRS is similar to that achieved when adding the PheRS to age and sex (**Supplement Figure 2A**).

**Supplement Figure 8**. In the meta-analysis PheRS had significantly ($p<0.05$) larger HRs per 1-SD for 3/13 diseases (epilepsy, MDD, lung cancer) and PGS significantly larger HRs for 6/13 diseases (colorectal cancer, breast cancer, prostate cancer, T2D, AF), for the other diseases there was no significant difference between the two predictors. Overall, we we found the largest association of the PGS for prostate cancer (meta-analyzed HR: 1.80, 95% CI: 1.75-1.85), T2D (meta-analyzed HR: 1.70, 95% CI: 1.64-1.77), and gout (meta-analyzed HR: 1.1.64, 95% CI: 1.59-1.69).

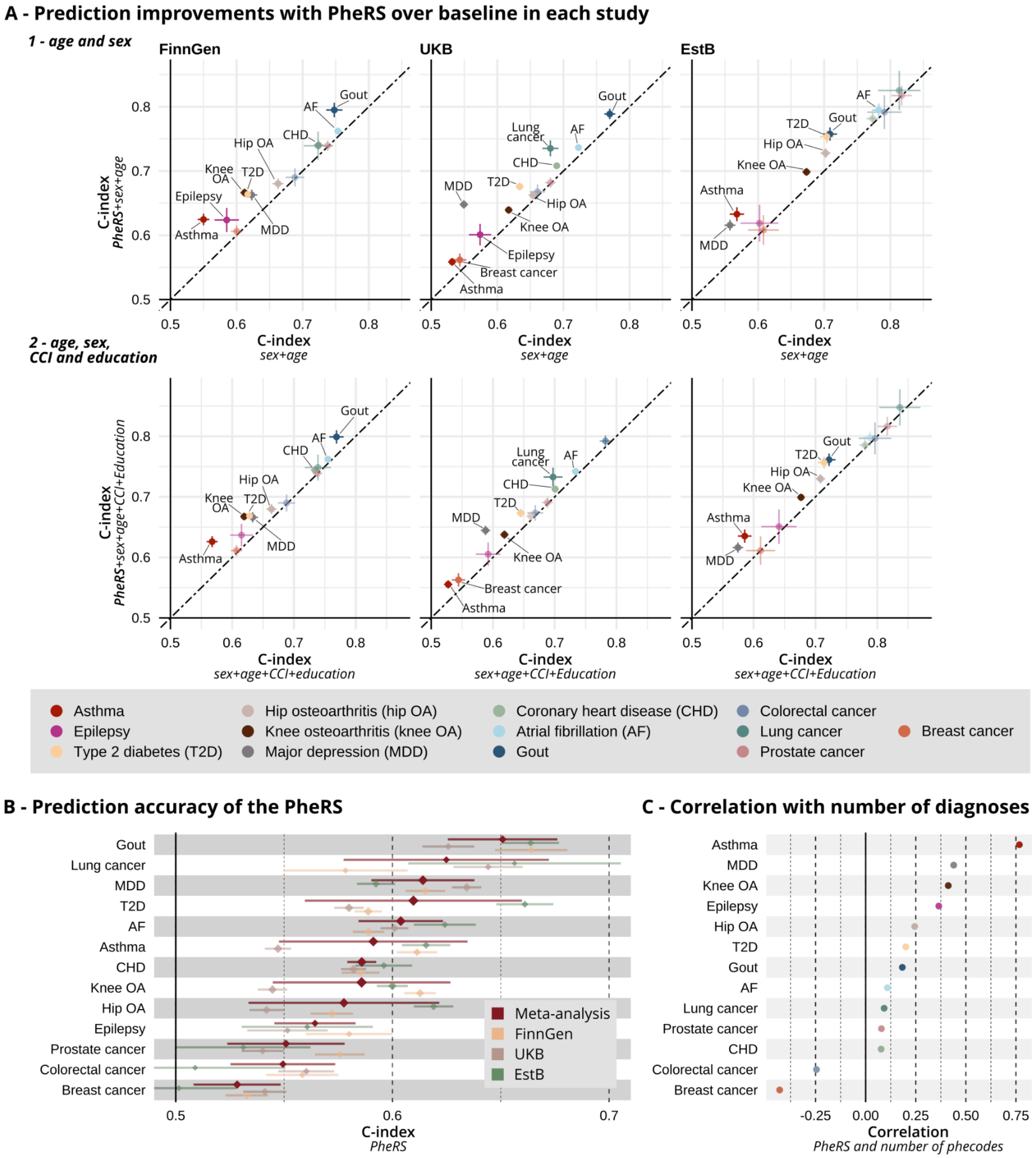


#### Supplement Figure 1

**Panel A: 1 -** Increase in prediction accuracy when adding the PheRS to a baseline model with age and sex in all three studies. The c-index and 95% CIs of the baseline model (x-axis), compared to a model with added PheRS (y-axis). Diseases with significant ($p<0.05$) increases in a study are labeled. **2 -** Increase in prediction accuracy when adding the PheRS to a baseline model with additional predictors education and CCI. Diseases with significant ($p<0.05$)

increases in a study are labeled. **Panel B:** The c-index of the PheRS model and 95% CIs separately in each study (FinnGen: yellow, UKB: brown, EstB: green) and meta-analyzed results (red). The effect of age and sex was removed by regressing them out from the PheRS. **Panel C**: Correlation between the PheRS and the total number of unique phecodes of each individual.

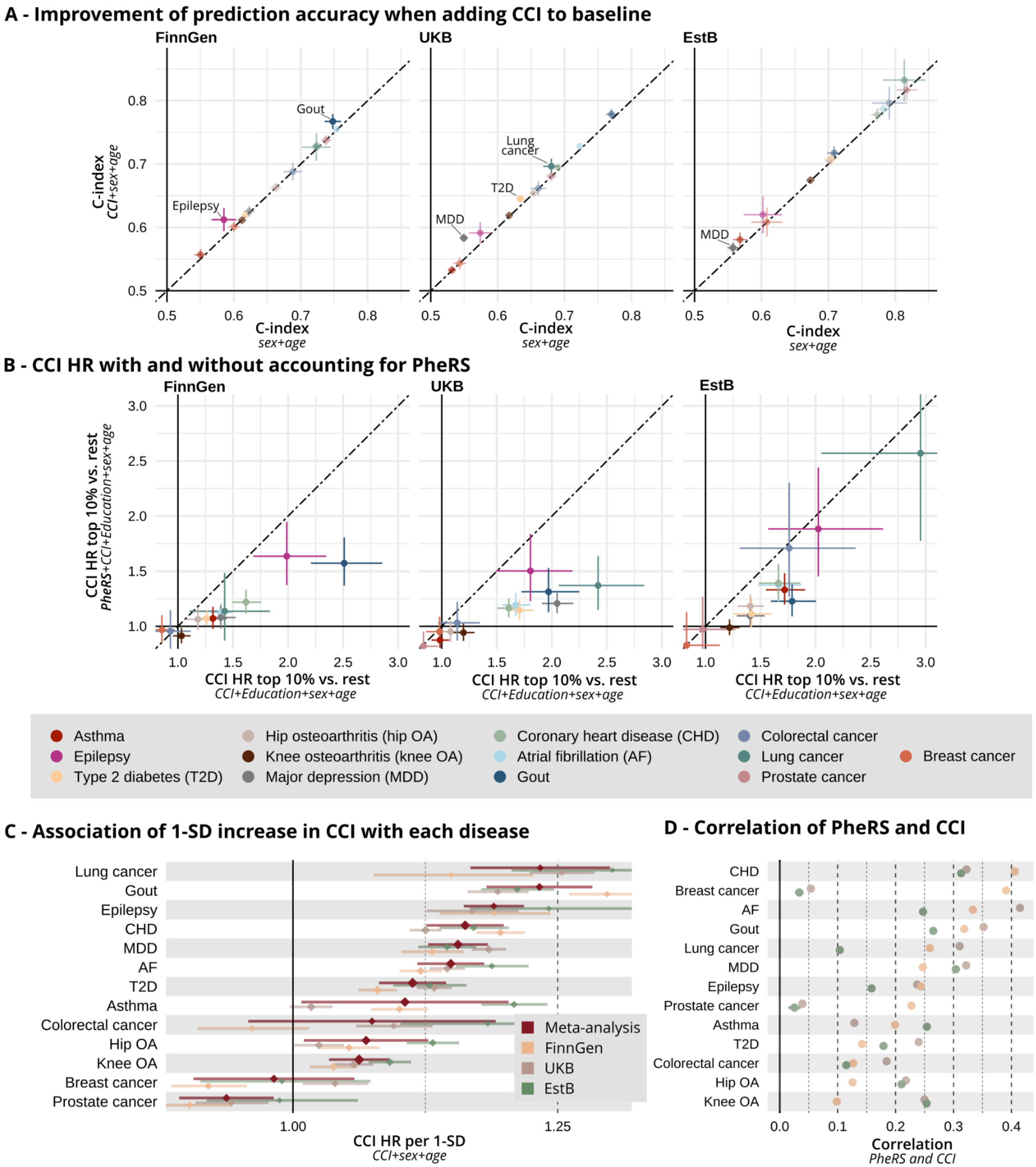


#### Supplement Figure 2

**Panel A:** Increase in prediction accuracy when adding CCI to a baseline model with age and sex in all three studies. The c-index and 95% CIs of the baseline model (x-axis), compared to a model with added predictors (y-axis). Diseases with significant (p<0.05) increases in a study are labeled. **Panel B**: Association of the CCI with each disease in a model without (x-axis) and with the PheRS (y-axis). The HRs are for the top 10% of individuals with the highest score compared to the rest 90%. Age and sex are regressed-out from the CCI. The top 10% corresponds largely to individuals with a CCI>=2 and a few younger ones with a CCI of 1. **Panel C:** Association of the CCI with each disease. This shows the increase in relative risk (HR and 95% CI) with 1-SD increase of the CCI. **Panel D**: Correlation of the PheRS and CCI after regressing-out the effect of age and sex.

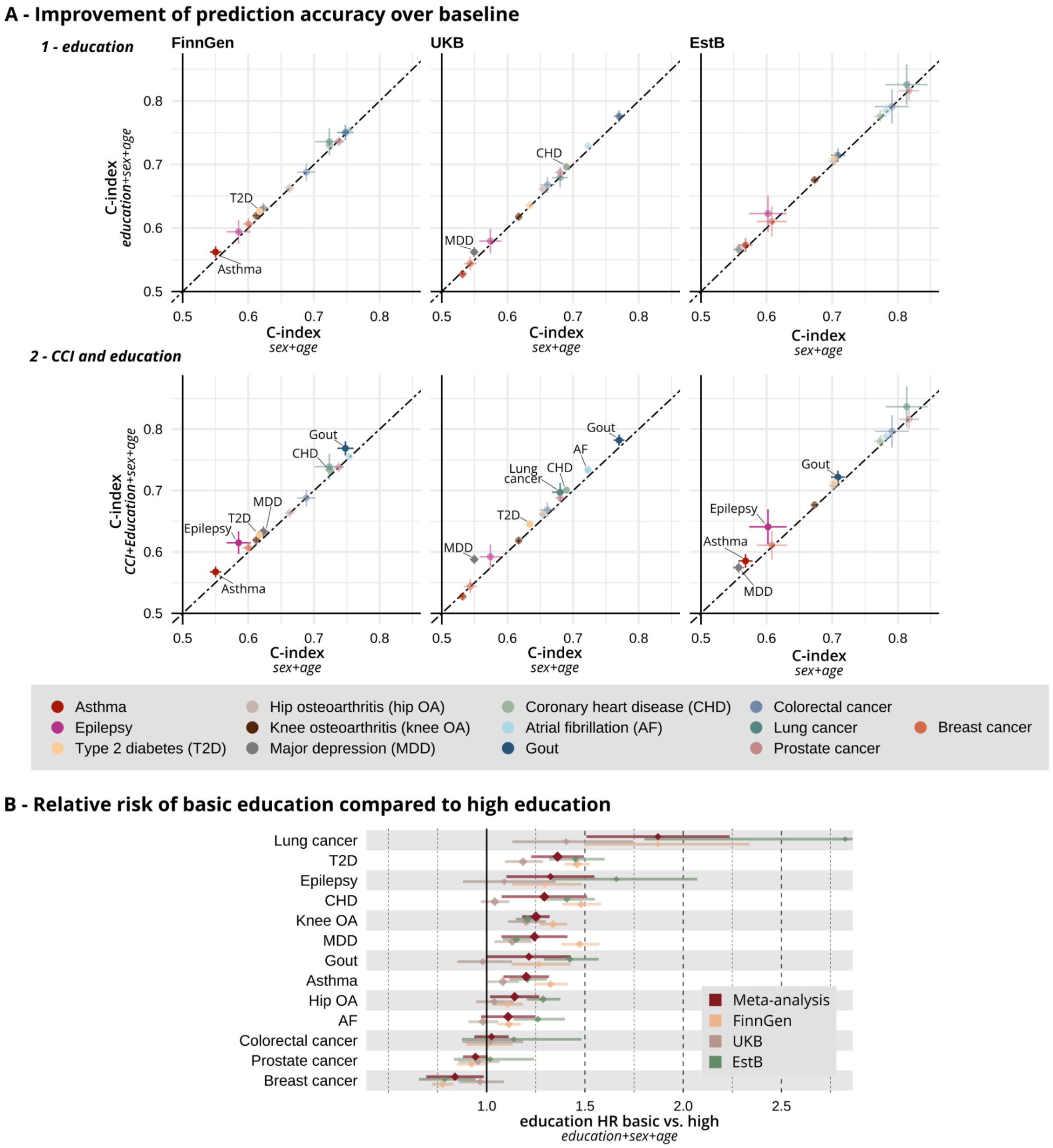


**Supplement Figure 3**

**Panel A:** Increase in prediction accuracy when adding education (**1**) and both education and CCI (**2**) to a baseline model with age and sex in all three studies. The c-index and 95% CIs of the baseline model (x-axis), compared to a model with added education (y-axis). Diseases with significant ($p<0.05$) increases in a study are labeled. **Panel B:** Association of lower education with each disease. Here we show the increase in relative risk (HR and 95% CI) for individuals with lower education (ISCED-11<5) compared to those with high achieved education level (ISCED-11>=5).

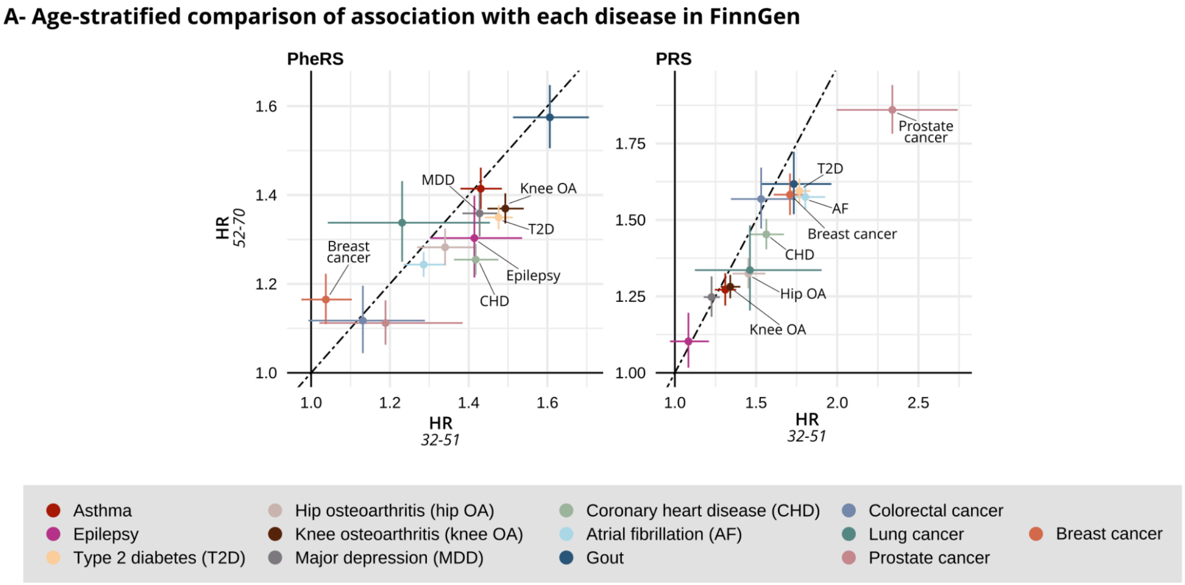


#### Supplement Figure 4

HRs of the PheRS in the younger group (32-51) compared to older individuals (52-70). The PheRS were trained on all individuals aged 32-70. Diseases with significant ($p<0.05$) differences in HRs between the groups are labeled.

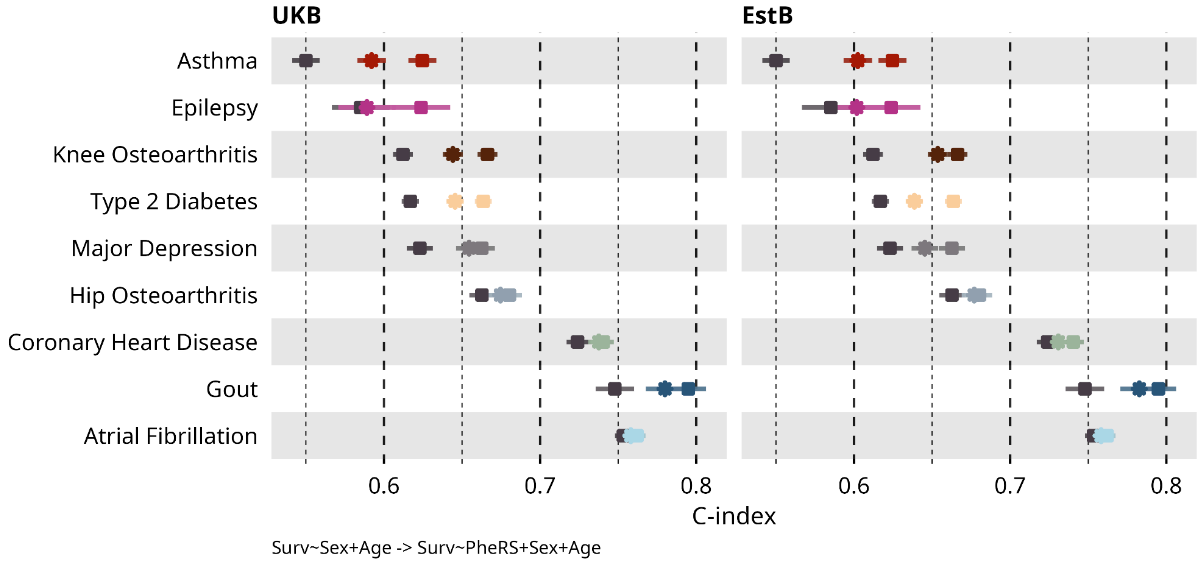

#### Supplement Figure 5

Improvement in predictive accuracy in FinnGen with FinnGen-trained and externally-trained PheRS models. The C-index and 95% CI of the baseline model with age and sex and the model with added PheRS. The c-index of the baseline models is shown by the dark grey boxes (the leftmost marker for each disease). The results of the PheRS models trained in FinnGen are shown as boxes with the color of each disease and the externally-trained PheRS as stars. The left panel shows the results with the UKB-trained and the right panel the EstB-trained models.

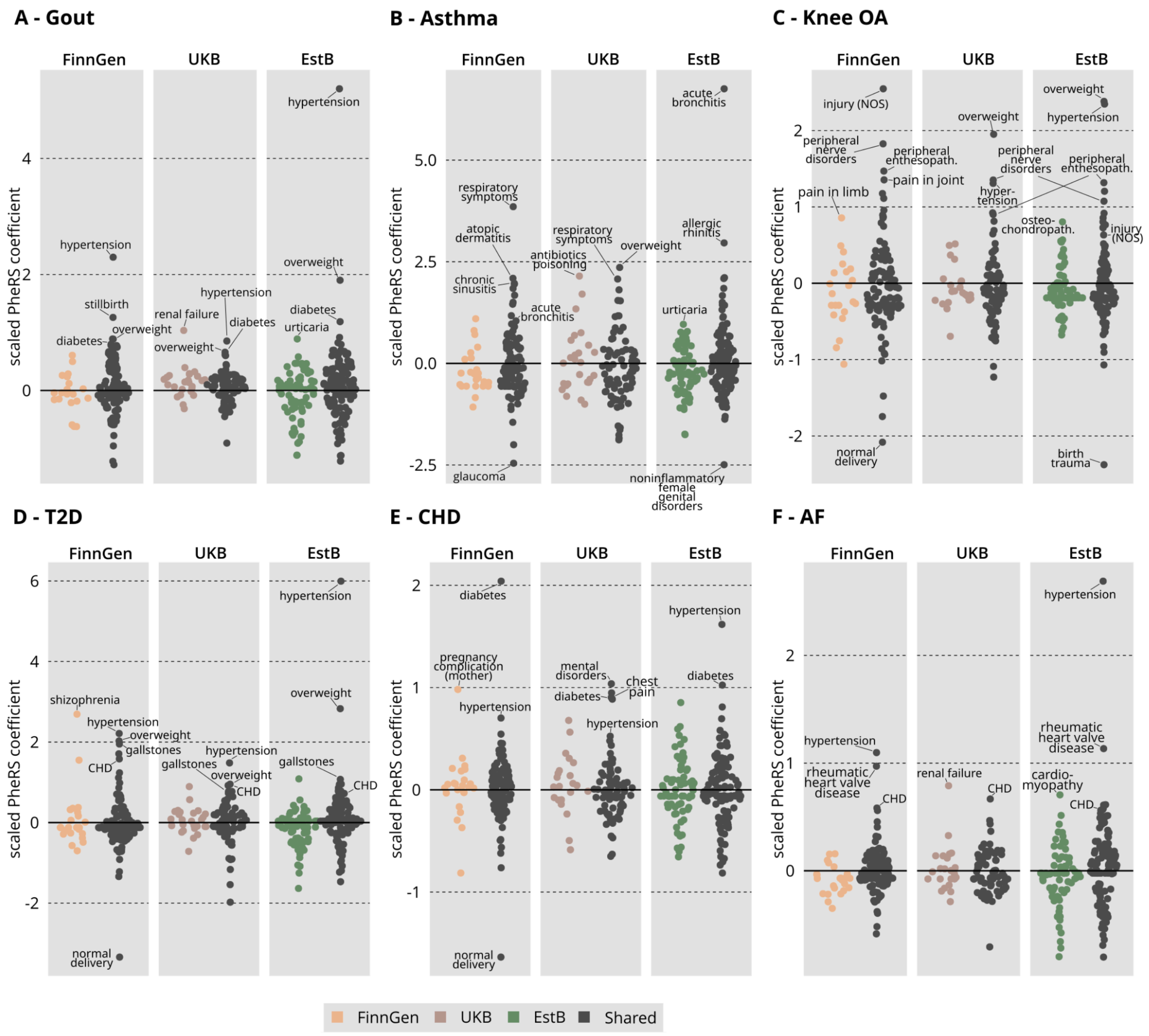


#### Supplement Figure 6

A detailed look on all the PheRS coefficients in the three studies for **Panel A:** Gout. **Panel B:** Asthma. **Panel C:** Knee osteoarthritis. **Panel D:** Type 2 diabetes. **Panel E:** Coronary heart disease. **Panel F:** Atrial fibrillation. Black color marks common phecodes in the PheRS models across the studies, while other colors indicate biobank-specific codes (yellow=FinnGen, brown=UKB, green=EstB). PheRS coefficients are standardized to 0 mean and 1 standard deviation for each model separately for easier comparison of coefficient importances across the studies.

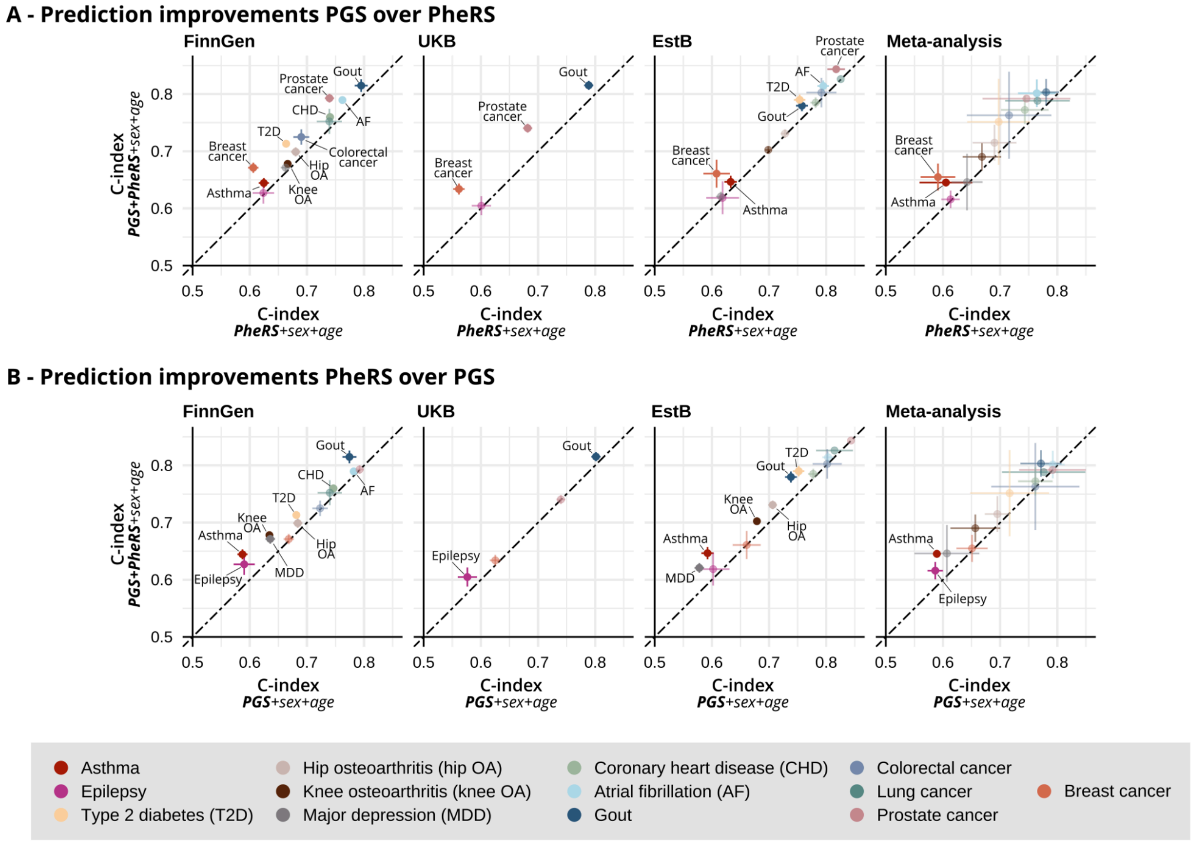


#### Supplement Figure 7

**Panel A:** Improvement of prediction discrimination when adding PGS to a model with PheRS, age, and sex. The c-indices and 95% CI of the PheRS+age+sex model (x-axis) and the model with PheRS+PGS+age+sex models (y-axis). Diseases with significant ($p<0.05$) increases in a study are labeled. **Panel B:** Improvement of prediction discrimination when adding PheRS to a model with PGS, age, and sex. The c-indices and 95% CI of the PGS+age+sex model (x-axis) and the model with PheRS+PGS+age+sex models (y-axis). Diseases with significant ($p<0.05$) increases in a study are labeled. Due to sample overlap with the GWASs, PGS could only be calculated for 4 diseases in UKB (see Methods for details).

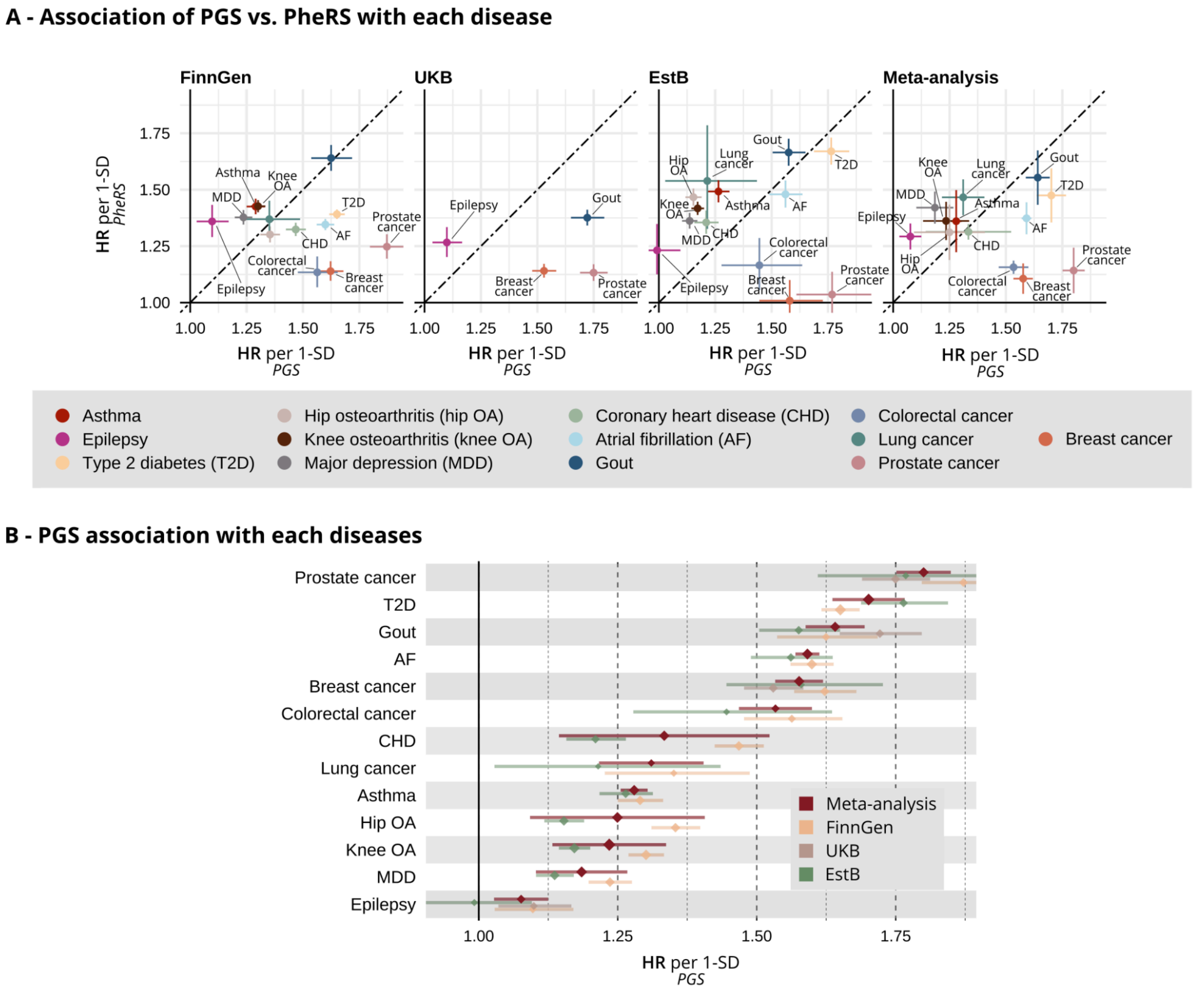


**Supplement Figure 8**

**Panel A**: Association of PGS (x-axis) vs. PheRS (y-axis) with each disease in each study. HRs and 95% CIs.The HRs are shown for 1-SD increase of the PheRS after regressing-out age, sex, and the first 10 PCs. Diseases with significant ($p<0.05$) differences in HRs between the groups are labeled. **Panel B:** Association between PheRS and disease onset during the prediction period. The HRs and 95% CIs in each study - FinnGen: yellow, UKB: brown, EstB: green - and meta-analyzed results (red). The HRs are shown for an increase of the PheRS by 1 standard deviation (SD) after regressing-out age, sex, and the 10 first PCs. Due to sample overlap with the GWASs, PGS could only be calculated for 4 diseases in UKB (see Methods for details).
